# Data-driven method to infer the seizure propagation patterns in an epileptic brain from intracranial electroencephalography

**DOI:** 10.1101/2020.07.30.20165183

**Authors:** Viktor Sip, Meysam Hashemi, Anirudh N. Vattikonda, Marmaduke M. Woodman, Huifang Wang, Julia Scholly, Samuel Medina Villalon, Maxime Guye, Fabrice Bartolomei, Viktor K. Jirsa

## Abstract

Surgical interventions in epileptic patients aimed at the removal of the epileptogenic zone have success rates at only 60-70%. This failure can be partly attributed to the insufficient spatial sampling by the implanted intracranial electrodes during the clinical evaluation, leading to an incomplete picture of spatio-temporal seizure organization in the regions that are not directly observed. Utilizing the partial observations of the seizure spreading through the brain network, complemented by the assumption that the epileptic seizures spread along the structural connections, we infer if and when are the unobserved regions recruited in the seizure. To this end we introduce a data-driven model of seizure recruitment and propagation across a weighted network, which we invert using the Bayesian inference framework. Using a leave-one-out cross-validation scheme on a cohort of fifty patients we demonstrate that the method can improve the predictions of the states of the unobserved regions compared to an empirical estimate. Furthermore, a comparison with the performed surgical resection and the surgery outcome indicates a link between the inferred excitable regions and the actual epileptogenic zone. The results emphasize the importance of the structural connectome in the large-scale spatio-temporal organization of epileptic seizures and introduce a novel way to integrate the patient-specific connectome and intracranial seizure recordings in a whole-brain computational model of seizure spread.

## 1. Introduction

A possible treatment for patients with drug-resistant epilepsy is a surgical intervention aimed at the removal of the suspected epileptogenic zone (EZ), i.e. the brain region responsible for the initiation of the seizures whose removal would result in seizure freedom. However, these surgical interventions have success rates in rendering the patients seizure-free at only 60-70% (Jehi et al., 2015; Baud et al., 2018). Why that can be? The current standard in pre-surgical evaluation is the use of either implanted depth electrodes (stereo-electroencephalography, SEEG) or subdural electrode grids (Jayakar et al., 2016). Interpreting the electrographic signals is however not straightforward due to the complex local dynamics and interactions between brain regions (Bartolomei et al., 2008, 2017), and the degree of epileptogenicity of brain structures and the extent of highly epileptogenic tissue might be misestimated. Furthermore, intracranial EEG does not allow for the exploration of the whole brain, and it is biased to the regions suspected to be part of the epileptogenic network based on the non-invasive evaluation. This introduces a risk that the highly epileptogenic tissue is not fully explored by the implantation, leading again to an incomplete resection.

In this work we explore if these issues can be solved by exploiting the role of structural connections in spatio-temporal seizure organization. In healthy brains there exists a strong link between the structural and functional connectivity (Ghosh et al., 2008; Honey et al., 2009). Studies indicate that in epileptic brains the structural connectivity is altered (Besson et al., 2017) as is the structure-function relationship (Wirsich et al., 2016), and, importantly for this study, that the long-range white matter connections shape the spread of epileptic seizures (Milton et al., 2007; Proix et al., 2017; Parker et al., 2018).

The increasing availability of diffusion-weighted magnetic resonance imaging in clinical practice allowed for building patient-specific structural brain networks, which opened the way for network-based computational models of epileptic activity (Taylor et al., 2014). Several such models appeared in past years with the aim to explore the possibilities of surgical interventions and to predict their outcome (Hutchings et al., 2015; Jirsa et al., 2017; Proix et al., 2017; An et al., 2019; Olmi et al., 2019). Importantly, unlike the models based on the functional connectivity networks derived from interictal and/or ictal intracranial EEG recordings (Goodfellow et al., 2016; Sinha et al., 2017; Lopes et al., 2017, 2018; Laiou et al., 2019), the models based on the structural connectivity are not spatially restricted to the implanted brain regions and can simulate the whole brain dynamics. In some of the models, the heterogenity of the node behavior is caused only by the underlying connectivity (Goodfellow et al., 2016; Sinha et al., 2017). In others a spatially heterogeneous parameter representing node excitability is introduced; these models explore the effects of the heterogenity of local parameters in small synthetic networks (Terry et al., 2012; Hebbink et al., 2017; Lopes et al., 2020), or model the whole-brain dynamics with the local excitability informed by the patient-specific anatomy (Hutchings et al., 2015) or by the clinical hypothesis of the node excitability (Jirsa et al., 2017; Proix et al., 2017; Olmi et al., 2019).

While these network models of epilepsy can provide valuable insight into the role of the network in seizure organization, they were designed with forward simulations in mind and might not pose an easy target for model inversion. Here by model inversion we mean the process of finding the model parameters (such as the parameters of the neural masses in network nodes or connection strengths) that can best explain given observations of the network dynamics. Indeed, most of the model inversion studies related to epileptic seizures so far dealt with a single neural mass, small networks of neural masses, or uncoupled neural masses (Murta et al., 2012; Freestone et al., 2014; Papadopoulou et al., 2015; Cooray et al., 2016; Papadopoulou et al., 2017; Rosch et al., 2018; Karoly et al., 2018). Only recently have studies on large-scale network model inversion appeared (Hashemi et al., 2020), however, inversion of large systems of coupled differential equations can still be prohibitively computationally expensive and requires a careful balance of the model parameterization, choice of the prior distributions of the parameters, and settings of the inversion method.

In this work we approach the problem of seizure propagation in a network from the other side, and we introduce a model of seizure propagation designed with the inversion in mind. As such, the model drops some complexity of the existing models while keeping the core elements of network models of seizure propagation: two possible states of a network node (either a healthy or a seizure state), the role of the network connections in seizure spread, and the role of heterogeneous node excitability. In exchange we obtain a model which can be reliably inverted both in a single-seizure regime to obtain seizure-specific regional excitabilities, or in multi-seizure regime to obtain the optimal hyperparameters of the model shared between seizures. The model is thus data-driven: we provide only its generic form and infer the parameters from the data. Using the Bayesian inference framework, we invert the model using the Markov Chain Monte Carlo (MCMC) method in order to quantify the uncertainty of the estimations.

In the conceptual view adopted here the brain network during a seizure is only incompletely observed, with some regions observed as non-seizing and some as seizing with a specific onset time of seizure activity (Fig. 1A). Using a collection of seizures recorded from several patients, patient-specific brain networks, and the introduced model, we perform the model inversion to obtain the hyperparameters of the model and the seizure-specific excitabilities, and to fill in the unknown onset times of hidden nodes (Fig. 1B). We validate the method on synthetic, model-generated data, as well as on real data recorded from patients with drug-resistant epilepsies. To do the latter we use two complementary approaches (Fig. 1C): The leave-one-out validation tests how well can the method predict the state and onset time of the hidden regions by excluding one observed region from the data, fitting the model without it, and comparing the prediction with the left-out information. The resection validation tests how well can the method predict the surgery outcome using the patient-specific fitted models and the actual resection performed.

**Figure 1:**
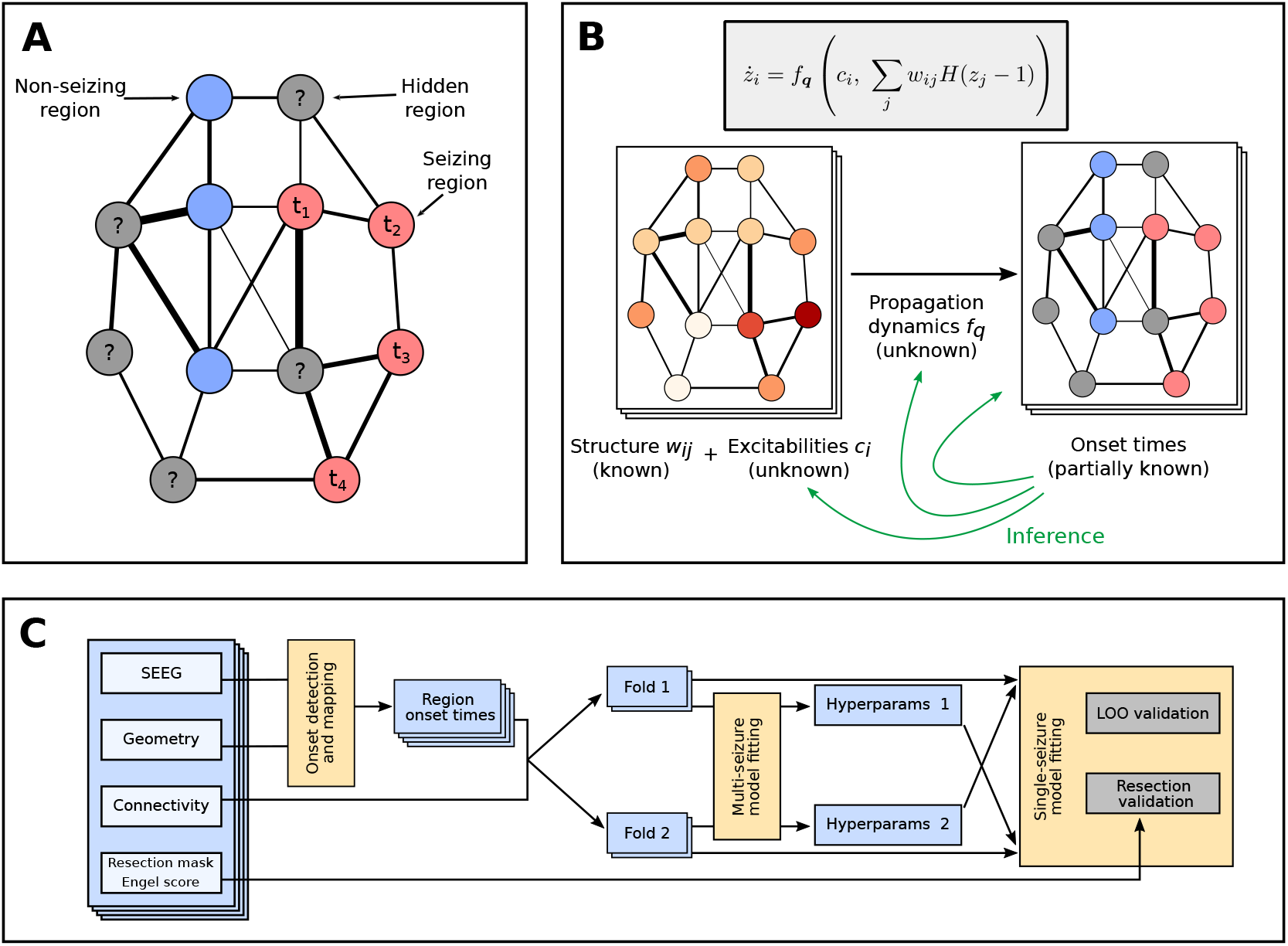
Main concepts and organization of the work. (A) Observation of a spreading seizure in the brain network. Due to the sparseness of the implanted electrodes, only some regions are observed; for those we know their state, non-seizing or seizing, and the onset time in the latter case. (B) Generative model of the seizure and the problem of the inference. We assume that the region onset times result from initially unknown propagation dynamics, shared among all seizures, and depend on the known patient-specific network structure and unknown seizure-specific region excitabilities. The goal of the inference is to infer the form of the propagation dynamics and the seizure-specific region excitabilites, and thus also the missing onset times. (C) The workflow used in this study for the validation on real data of fifty subjects. From the SEEG recordings the channel onset times were extracted and mapped onto brain regions. The data set was then divided into two folds, and each was fitted separately with the multi-seizure model to infer the model hyperparameters. The leave-one-out validation and the resection validation were then performed using the single-seizure model with hyperparameters obtained from the other fold.

## 2. Results

### 2.1. Overview of the method

While the detailed description of the method is given in Materials and Methods, here we provide a general overview of the method and of the assumptions behind it. At the core of the method is the dynamical model of seizure propagation in the brain network. The model is constructed so it, in the simplest fashion possible, expresses the following assumptions about the seizure propagation:

- The seizure propagates through the brain network, represented as a weighted network *W* = {*w*_*ij*_} *∈* ℝ^*n*×*n*^, where *n* stands for the number of brain regions.
- Each node of the network is at any given point in time either in a healthy state or in a seizure state.
- The sudden transition of a brain region *i* from the healthy to the seizure state is driven by continuous changes in the slow variable *z*_*i*_, loosely corresponding to the slow permittivity variable in the Epileptor model (Jirsa et al., 2014) or to the usage-dependent exhaustion of inhibition in the model of Liou et al. (2020). All regions are initially in a healthy state, and any region starts to seize when its slow variable crosses a given threshold.
- The rate of change of the slow variable of a region *i* depends only on its inner excitability *c*_*i*_ and the input it receives from the connected seizing regions.

Formally, the dynamical model of the seizure propagation is written as follows:

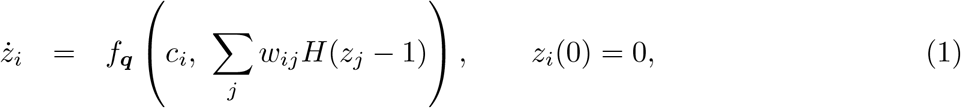

where *z*_*i*_ is the slow variable, *c*_*i*_ is the region excitability, *w*_*ij*_ are the connection strengths, and *H*(*z*_*j*_ − 1) is the Heaviside function, representing the seizure threshold at *z*_*j*_ = 1. We call the time point *t*_*i*_ when region *i* crosses the threshold and starts to seize at *onset time* of region *i*. Finally, the function *f*_***q***_ is the *excitation* function parameterized by the parameter vector ***q***.

We require the function *f*_***q***_ to be positive; consequently, all regions will switch to the seizure state in finite time. That is clearly not the case in reality, and we thus associate all regions that start to seize after limit time *t*_lim_ with non-seizing regions. This constant expresses the time scale we consider relevant for seizure spread; in this work we set *t*_lim_ = 90 s. Associating late-seizing with non-seizing regions might at first seem like a poor approximation of reality, it however serves a crucial purpose for the model inversion: it allows us to avoid a problem of a discrete nature (inferring *if* a region seizes) and replace it with a continuous problem (inferring *when* a region starts to seize), which is better suited for inference using the MCMC methods. This approach can be justified also from the clinical perspective: the primary objective of the method is to understand the relations between the regions that seize early, and the interactions of the late-seizing or non-seizing regions are only of secondary importance.

Given the parameters ***q***, the connectome matrix *W*, and the excitabilities ***c***, the dynamical model determines the onset times ***t*** (Fig. 2). The problem we are facing is however the opposite: given the partially observed onset times and the connectome matrices (for multiple seizures and multiple subjects) we want to infer the parameters ***q*** of the excitation function, which we assume are shared among all seizures, and the seizure-specific excitabilities. Once these parameters are inferred, the unknown onset times can be easily calculated. We perform this inversion in Bayesian inference framework using the statistical model built upon the dynamical model with additional assumptions:

**Figure 2:**
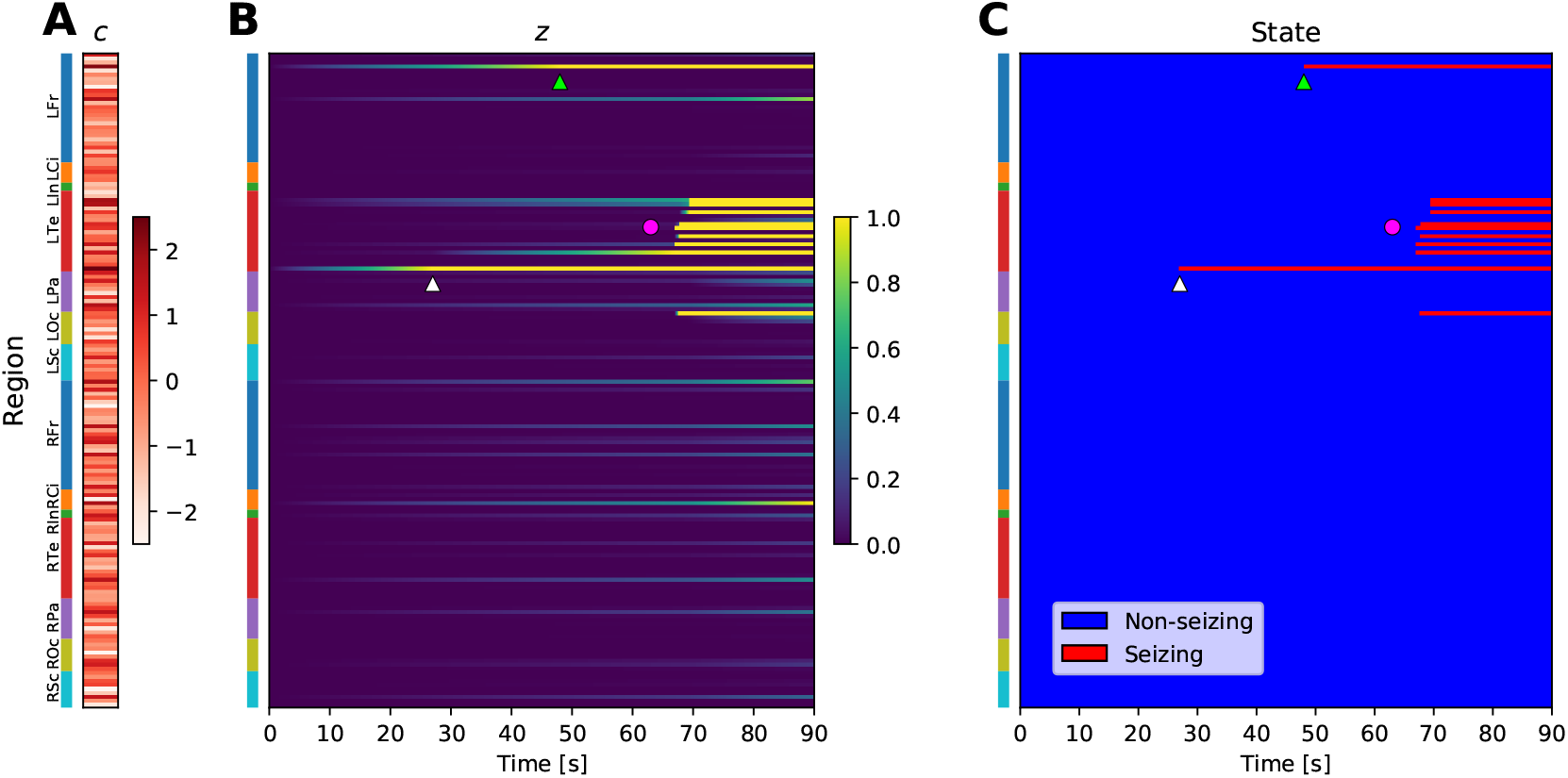
Example of a simulated seizure in real brain network. (A) The region excitabilities *c* are randomly sampled from a standard normal distribution. Brain regions are located on the vertical axis, colored bars indicate the anatomical grouping of the regions. (B) Slow variable *z*, simulated using the sampled excitabilities and *strong coupling* excitation function (see Fig. S2) on a brain network of subject 1. (C) Seizure state of the regions, obtained by thresholding the slow variable at *z* = 1. Due to the elevated excitability of the regions, the seizure starts in the Rhinal cortex in left temporal lobe (white triangle), and with some delay in Pars orbitalis in left frontal lobe (light green triangle). Eventually, large portion of the left temporal lobe is recruited (magenta circle). Abbreviations: Lxx/Rxx - left/right hemisphere, Fr - frontal lobe, Ci - cingulate cortex, In - Insula, Te - temporal lobe, Pa - parietal lobe, Oc - occipital lobe, Sc - subcortical structures. See Supplementary Table S2 for a full list of regions.

- The propagation dynamics (or, in the model terms, the parameters ***q*** of the excitation function) is shared among all seizures. The individual variability in the observed onset times is therefore assumed to be caused only by the seizure-specific region excitabilities ***c*** and the patient-specific structural network *W*.
- The excitabilities ***c*** are *a priori* assumed to follow the standard normal distribution *N* (0, 1).
- The onset times are detected inexactly with normally distributed observation error.

These assumptions lead to a hierarchical statistical model: the hyperparameters at the top level are the parameters ***q*** shared among all seizures, while at the bottom level are the seizure-specific excitabilities ***c***. Intuitively, the goal of the inference is to find such parameterization ***q*** of the excitation function so that the observations of all seizures can be explained by excitabilities that are as close to being normally distributed as possible given the constraints posed by the model formulation. The model, as we will demonstrate, can be robustly inverted even for large brain networks. Furthermore, due to its simplicity, mathematical statements about the existence and uniqueness of the solution can be made in case of no observation error (SI Text).

### 2.2. Onset time detection and mapping

First step in applying the model to real patient data is the detection of the onset times of seizure activity in the recorded SEEG signals (Fig. 3A) and the subsequent mapping of the channel onset times to the brain regions defined by the brain parcellation (Fig. 3B). In our cohort of fifty patients, there was between ten to thirty-five regions observed (i.e. with at least one assigned channel) out of 162 regions in the brain parcellation (Fig. 3C); these observed regions have predominantly less than four channels assigned (Fig. 3D). The channel to region mapping may lead to one region having multiple channels assigned with different seizing/non-seizing status or different onset times. Such occurrence indicates that the parcellation is not sufficiently fine to properly capture the spatio-temporal dynamics of the seizure, however, we observe that the state of over 90% of the observed regions is unambiguous (Fig. 3E). Furthermore, the difference on the earliest and latest assigned onset time is below ten seconds for 80% of the seizing regions (Fig. 3F).

**Figure 3:**
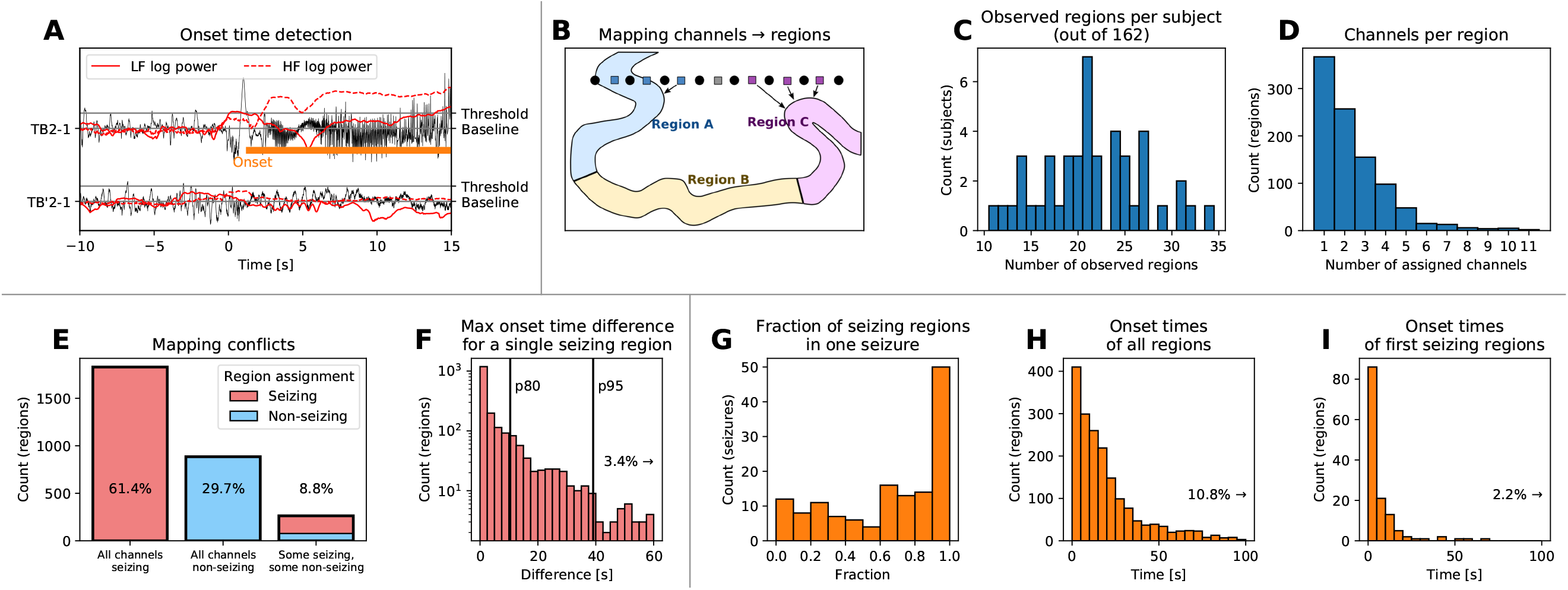
Onset time detection and mapping to brain regions. (A) Example of the onset time detection. The onset time on a bipolar SEEG channel is detected by computing the power in lower and high frequencies, normalizing it to preictal levels, and detecting when the power increases above a given threshold. This results in marking a channel as seizing (with detected onset time, upper trace) or non-seizing (lower trace). (B) Channels are assigned to brain regions based on their physical distance. If multiple channels are assigned to the same region, the seizing or non-seizing status is determined based on the majority of channels. If the region is seizing, the region onset time is defined as the median of all assigned onset times, taking the non-seizing regions into account as well with onset time equal to infinity. (C) Histogram of observed regions among all patients in the study. (D) Histogram of assigned SEEG channels per observed region. (E) Histogram of regions based on the seizing state of the assigned channels, indicating where the channel to region mapping leads to a conflict in the region seizing state. Ideally, there would be no regions with some seizing and some non-seizing assigned channels. (F) Histogram of differences between the earliest and latest onset time of assigned channels. Vertical lines indicate the 80th and 95th percentile. (G-H) Results of the detection and mapping. (G) Histogram of the fractions of the seizing regions for all seizures. (H) Detected onset times of all seizing regions, relative to the clinically marked seizure onset. (I) Detected onset times of the first seizing region of every seizure, relative to the clinically marked seizure onset.

Fig. 3G-I shows the final results of the detection and mapping procedure on the patient data. Seizures where very few, some, or all of the observed regions are seizing are present (Fig. 3G); seizures where close to all observed regions are seizing are more represented, possibly reflecting the bias in the SEEG implantation towards the regions where the seizure activity is expected. The frequency of occurrence of detected onset times decays with increasing delay from the clinically marked seizure onset (Fig. 3H). In the majority of the seizures, the earliest seizure onset follows the clinically marked seizure onset with little delay (Fig. 3I).

### 2.3. Hyperparameter learning

Next, we used the cohort data to infer the hyperparameters of the model. To avoid a data reuse in the subsequent leave-one-out validation, we divided the cohort into two folds of equal size, and the multi-seizure model was fitted twice, independently for each fold. That led to two sets of estimated parameters. At most two seizures per single patient were used for the fitting to avoid biasing the model towards the patients with more recorded seizures.

Fig. 4A shows the inferred posterior distribution for the model hyperparameters, together with two measures of convergence: split-chain scale reduction factor 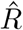 and number of effective samples *N*_*eff*_ (Gelman et al., 2013). In some cases the diagnostic values lie slightly outside of the generally recommended range 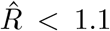 < 1.1 indicating imperfect mixing of the MCMC chains; we consider it acceptable given the practical limitations of the required time for the computations and considering that the hyperparameter posterior distribution will be reduced to point estimate for the second step of fitting the individual seizures.

**Figure 4:**
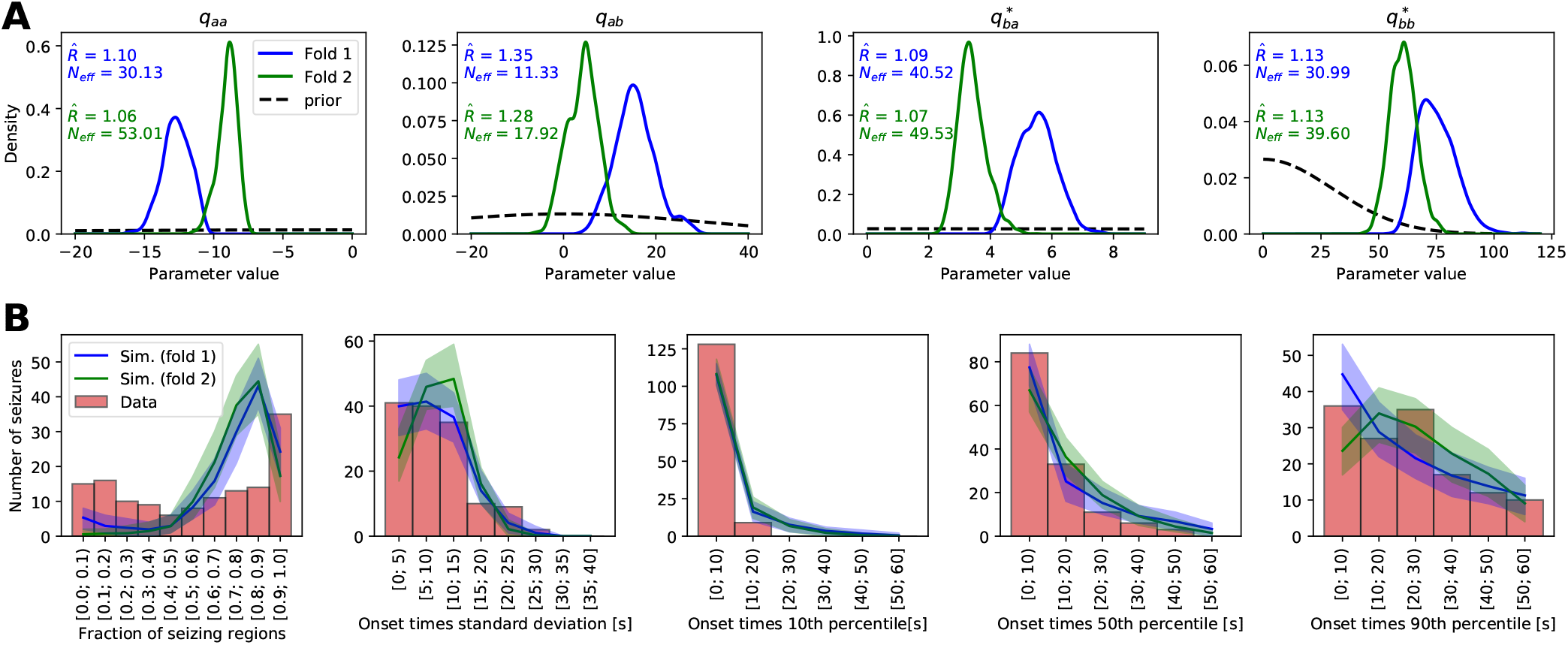
Results of the hyperparameter learning and posterior predictive checks. (A) Posterior (full lines) and prior (black dashed lines) distributions of the hyperparameters for the models fitted with two data folds. The text shows the split-chain scale reduction factor 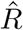 and number of effective samples *N*_*eff*_. Note the different x and y ranges of the panels. (B) Results of the posterior predictive checks with the fitted models. In all panels, the red histogram shows the properties of the real seizure ensemble, while the solid lines show the mean of the hundred ensembles of simulated seizures and the shaded areas indicate the 5 to 95 percentile range. The panels show, left to right, distribution of the fraction of the seizing regions in one seizure, standard deviation of the onset times of seizing regions, and 10th, 50th, and 90th percentile of the onset times of seizing regions.

Even though the inferred parameter posterior distribution for both folds do not overlap perfectly, they lie in the same region of parameter space (Fig. 4A and Fig. S5). The two folds contain data from entirely different patients, these results thus indicate that some common features of the seizures are indeed extracted from the data and that the models are not overfitted to any specific data set.

If the model is sufficiently flexible and fitted well, it should be able to generate synthetic seizures resembling the real ones. Testing if it does is the goal of posterior predictive checks. We took the two hyperparameter sets obtained by fitting the two folds, we randomly drew the excitabilities from the prior standard normal distribution, and we simulated an ensemble of 137 seizures - same number as of the real seizures in the data set. We repeated this process one hundred times to obtain hundred ensembles of simulated seizures for both folds and then we compared the statistical properties of the synthetic seizure ensembles with the ensemble of real seizures (Fig. 4B). Several observations can be made from the results. First, the two models from two folds produce statistically similar seizures, indicating again that some common features of seizure dynamics were extracted from the data and the models were not overfitted. Second, the models generate less seizures with only few regions recruited, while overpredicting the number of seizures with majority (but not all) of the regions recruited. The statistics of the onset times of seizing regions are however well reproduced.

### 2.4. Single-seizure inference examples

We present four examples of the inference results to illustrate the working of the method. Fig. 5 depicts a seizure where the inferred extent of recruited regions is spatially restricted and where the existence of hidden epileptogenic zone is predicted. In this specific case the resective surgery did not result in seizure freedom, it is thus possible that the epileptogenic zone was indeed not explored by the implantation. In contrast, Fig. S6 shows a temporal lobe seizure where the inference results mirror the observations, i.e. no early involvement of any non-observed regions is predicted, and the inferred epileptognic regions coincide with the observed early seizing regions.

**Figure 5:**
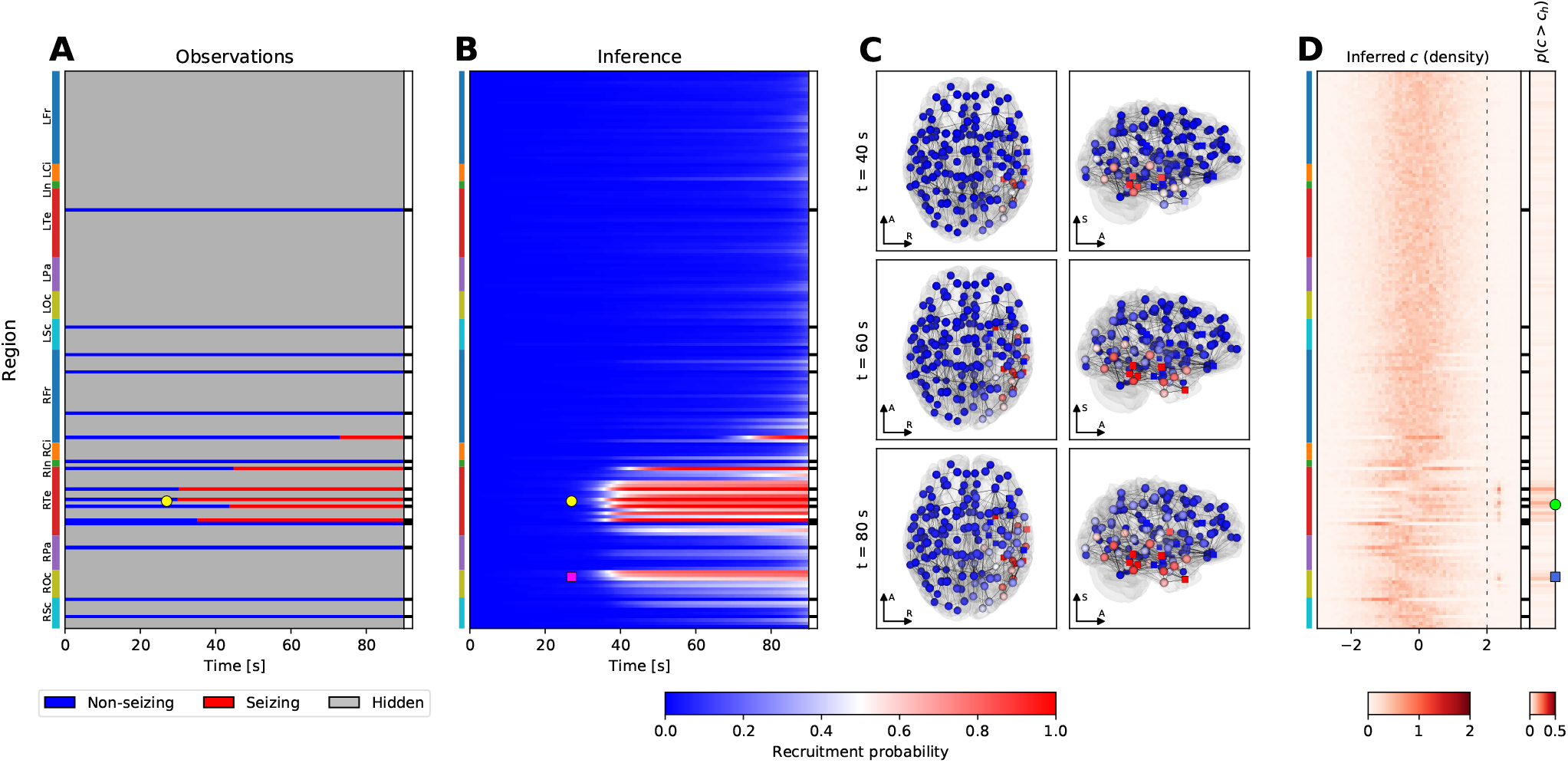
Example of the inference results on a seizure from subject 33. (A) Observation of the seizure. On vertical axis are the brain regions with color bars on the left indicating the anatomical grouping (abbreviations as in Fig. 2). On the horizontal axis is the time; the onset time of the first observed seizing region is always aligned to *t* = 30*s*. The black and white column on the right shows which regions were observed. (B) Results of the inference. The partial observations from panel A are completed by the inference; the plot shows the recruitment probability *r*_*i*_(*t*) = *p*(*t*_*i*_ ≤ *t*), i.e. the posterior probability that a region *i* is recruited at time *t*. In the observed regions the inferred probabilities follow closely the observations with some blurring around the onset due to the assumed observation noise. (C) Snapshots of the recruitment probabilities at three time points. Color code same as in panel B. The spheres and the cubes represent the hidden and observed regions respectively. Only top three percent of the strongest connections are shown for visual clarity, their thickness is proportional to the maximum of the two oriented connection strengths. Axes notation: R, Left-Right axis; A, Posterior-Anterior axis; S, Inferior-Superior axis. (D) Inferred excitability. Left subpanel shows the posterior distributions of the excitabilities, dashed line indicates the threshold of high excitability at *c*_*h*_ = 2. Right subpanel shows the probability of high excitability *p*(*c* > *c*_*h*_). (A-D) The seizure is observed to start and remain restricted in the right temporal lobe (yellow circle, panels A, B). The result of the inference in addition points to the involvement of several regions in the right occipital lobe (magenta square, panel B). The regions identified by the inference as possibly epileptogenic are mainly located in the right temporal lobe (green circle, panel D), however, the inference also points to a possible epileptogenic zone in the right occipital lobe (blue square, panel D).

An example of a more spatially extended seizure is shown on Fig. S7. The seizure is inferred to start in the right hippocampus before spreading to the majority of the right hemisphere and, eventually, also to the left hemisphere. Despite the inferred involvement of large portion of the brain, the inference points clearly to suspected epileptogenic zone due to the observed early involvement of the right hippocampus.

Finally, Fig. S8 shows often observed case of a seizure where a large majority of the regions is inferred to be recruited in the seizure at approximately same time. None of the regions is strongly inferred as epileptogenic, since it is difficult to identify which regions are the leaders and which are the followers of the seizure activity when the temporal delays of the recruitment are this small.

### 2.5. Leave-One-Out cross-validation

The goal of the Leave-One-Out (LOO) cross-validation is to assess whether we can correctly predict the seizure state and the onset times of the hidden regions. By definition, the information about the hidden nodes is not available, thus we cannot evaluate this directly. Instead, we adopt the LOO approach: For every seizure and every observed region we fit the single-seizure model to the data with the observation of that specific region left out. Then we can compare how well the prediction matches the left-out observation. To avoid reusing the same data twice - once for the fitting of the multi-seizure model and once for the LOO fitting - all seizures from one fold are fitted using the hyperparameters obtained from the other fold.

In total, the 3027 inferences of the single-seizure model were run, each with two MCMC chains. From these, in 179 cases one of the chains was stuck and in 28 both chains were stuck; the latter cases were excluded from further analysis. In the remaining results, 99.80% of regional excitability parameters had values of split chain reduction factors 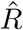 below 1.1 and effective sample size *N*_*eff*_above 30, indicating acceptable convergence for vast majority of the parameters.

The predictive power of the method was evaluated using the state prediction accuracy, quantifying the ability to predict the seizing or non-seizing state of a hidden region, and the onset prediction accuracy, quantifying the ability to correctly predict the exact onset time of a hidden region. To provide a baseline, the inference method was compared with two simpler approaches: an estimation based only on the onset times in other regions, and a weighted estimation based on the onset times in other regions and on the network structure. The results demonstrate high predictive power of the inference method (median state prediction accuracy 80.5%, median onset prediction accuracy 29.9%; Fig. 6A,D), yet it is not better than that of both the unweighted and the weighted estimates on group level (Fig. 6B,E).

**Figure 6:**
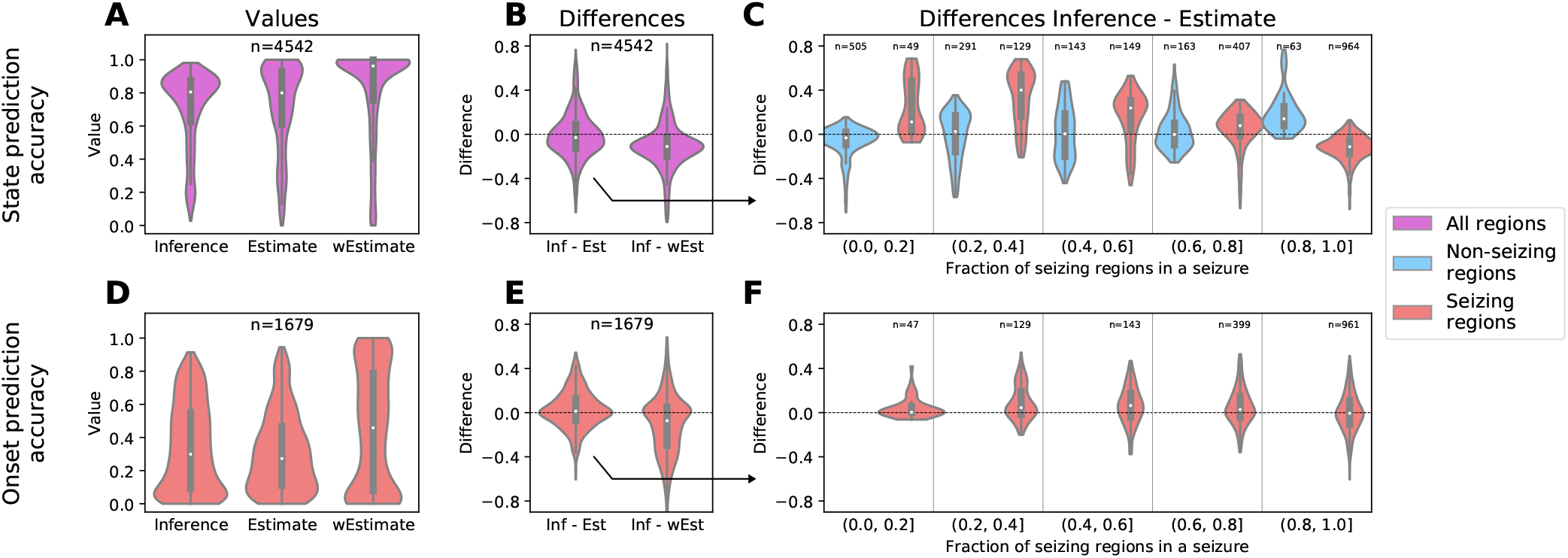
Results of the leave-one-out validation. (A-C) Results for the state prediction accuracy for all regions. (D-F) Results for the onset prediction accuracy for seizing regions. (A,D) Distribution of the state/onset prediction accuracy for the left-out regions obtained with the inference, estimate, and weighted estimate methods. Zero indicates wrong prediction, one is a perfect prediction. (B,E) Pair-wise differences of the accuracy obtained from the inference method and both estimates. (C,F) Differences of the accuracy of the inference and unweighted estimate, separated for the seizing and non-seizing nodes, and for the seizures with different fraction of seizing regions. In all panels, the violin plots show a kernel density estimate of a given variable. The inner boxplots show the median (white dot), interquartile range (IQR, gray bar) and adjacent values (upper/lower quartile +/−1.5 IQR, gray line).

Detailed analysis however reveals a finer structure in the results. We evaluated the prediction accuracies evaluated separately for seizures with different numbers of recruited regions, and furthermore separated the seizing and non-seizing regions (Fig. 6C,F). The results show a robust improvement obtained by the inference over the unweighted estimate for the seizing regions in seizures where the majority of the observed regions do not seize, especially for state prediction accuracy (Fig. 6C). In other words, the inference is better than the unweighted estimate in finding the hidden seizing regions in seizures where majority of the observed regions are not recruited, such as the seizure depicted on Fig. 5. This improvement is however not observed when compared to the weighted estimate (Fig. S9).

### 2.6. Validation against the resected regions

In addition to the predictions of the onset times in hidden regions, the product of the inference is also a spatial map of the excitability parameter *c* for every seizure. We evaluated whether this excitability is useful for localizing the epileptogenic zone. No ground truth to directly validate against exists, therefore we employed the following methodology: we restricted ourselves only to the patients that were operated and for which the post-operative MRI was available. We extracted which brain regions were resected, and using the inferred excitabilities for a specific patient and a specific seizure we performed an *in silico* resection, that is, we removed the resected regions from the network model and we performed the forward simulations with the excitabilities inferred from the observed seizures (Fig. 7A-C). The virtual resection was rarely sufficient to stop the seizures, indeed, large amount of brain regions continued seizing after the virtual surgery in patients of all Engel classes (Fig. 7D, top panel). However, the resection was more successful in relative reduction of the number seizing regions among the patients with Engel score I and II than among Engel III and IV patients (Mann-Whitney *U* = 67, *n*_*I,II*_ = 10, *n*_*III,IV*_ = 8, *p* = 0.019; Fig. 7D, bottom panel). Here by relative reduction we mean the number of regions where seizure activity was suppressed over the number of initially seizing regions. This result can be explained by the better match between the predicted epileptogenic and resected regions as illustrated by the precision-recall curve (Fig. 7E). In this context, the precision expresses how many of the predicted epileptogenic regions were resected, and the recall expresses how many of the resected regions were predicted to be epileptogenic.

**Figure 7:**
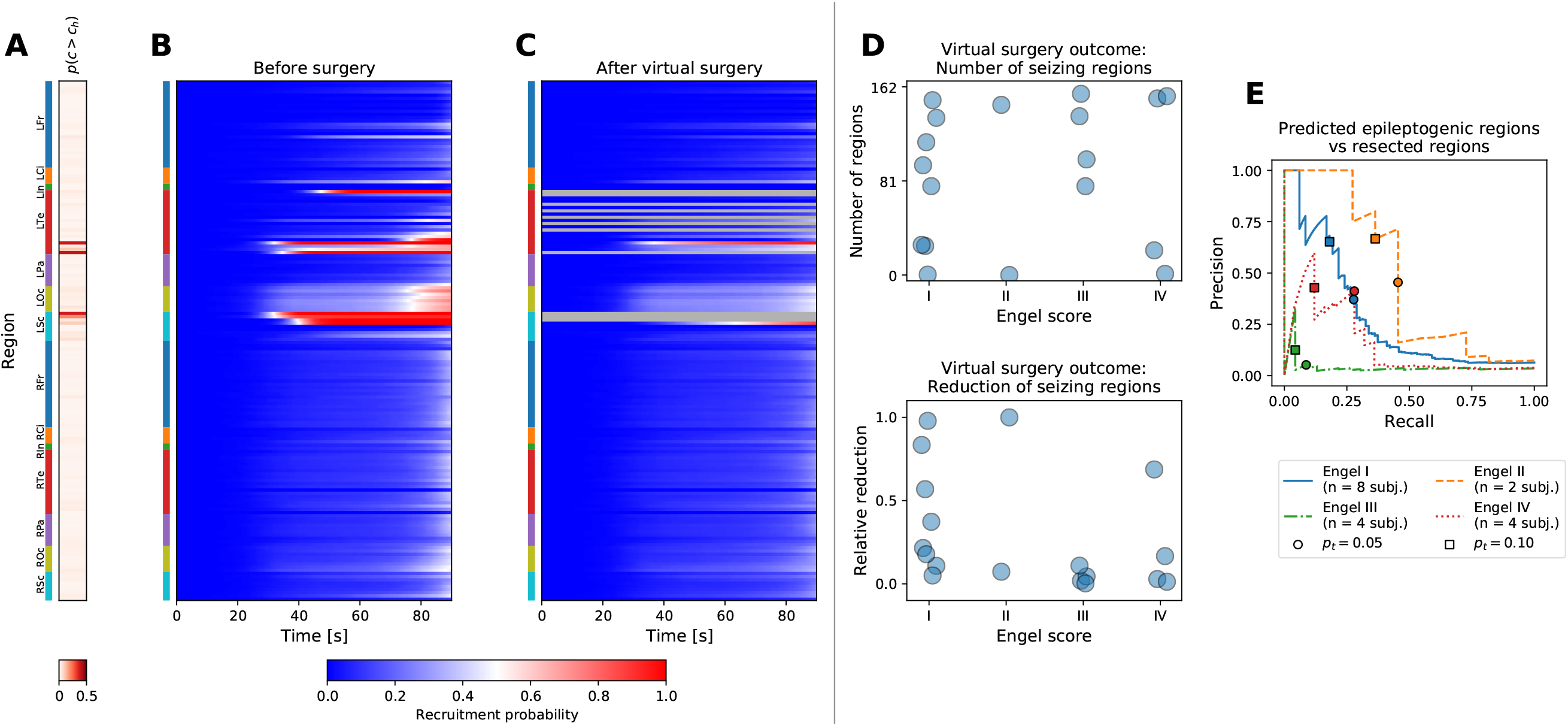
Virtual resection. (A-C) Example of a virtual resection on a seizure from patient 17, cf. Fig S6. (A) Inferred probability of high excitability. Anatomical abbreviations as in Fig. 2. (B) Pre-operative seizure dynamics as inferred from the data. (C) Post-operative seizure dynamics. The resected regions were removed from the model, and the dynamics was simulated using the excitabilities inferred from the pre-operative observations. The seizure activity is not completely stopped, but the number of seizing regions is reduced. (D) Outcome of virtual surgeries on a group level. Top panel shows the number of post-operative seizing regions *n*_postop_ (i.e. the regions with recruitment probability above 50% at *t* = *t*_lim_), bottom panel shows the relative reduction of the seizing regions compared to the pre-operative level, (*n*_preop_ − *n*_postop_)*/n*_preop_. For patients where multiple seizures were available, the values were averaged across seizures. (E) Precision-recall curves for evaluating the match between the performed resection and the inferred epileptogenicity. The precision and recall values were calculated for varying threshold *p*_*t*_ on high epileptogenicity, *p*(*c* > *c*_*h*_) > *p*_*t*_; the threshold *c*_*h*_ = 2 was kept constant.

## 3. Discussion

### 3.1. Main results

In this work we have developed and evaluated a novel method to infer the seizure propagation patterns in partially observed brain networks. The method is transparent in its assumptions and it is data-driven, meaning that the model hyperparameters are inferred from the patient cohort data, and only few clearly interpretable parameters need to be set by hand. It performs reliably when applied to synthetic data generated by the same model, although with limits caused by the incomplete observations of the seizures (Fig. S3). When predicting the states and onset times of the hidden regions in real data, the method performs better than the empirical estimate that does not use the network structure, but not better than the weighted estimate using the network structure (Fig. 6). While this result may be initially viewed with disappointment, it is important to keep in mind that unlike the weighted estimate our method provides a generative model which can be interrogated and its assumption can be questioned and modified. It is thus amenable to further development which may lead to more precise predictions. Furthermore, it provides the estimate of regional excitabilities. Indeed, the comparison with surgically resected regions and the surgery outcome indicates a link between the inferred excitabilities and the epileptogenic zone in the analyzed patients (Fig. 7).

### 3.2. Role of the large-scale structural connections in seizure propagation

Focal seizures are known to spread both locally and distally, but while recent experiments using *in vivo* rodent models shed more light on the mechanism of the long-range propagation (Rossi et al., 2017; Liou et al., 2018), it’s role in the large-scale spatio-temporal organization of epileptic seizures in human patients is not yet sufficiently understood. Here we have shown that a method based on the principles of network propagation predicts the states of the hidden regions better than an empirical estimate that does not use the network structure, but not better than an empirical estimate utilizing the network structure. Such results confirm the core tenet of our method that seizures spread along the long-range structural connections (Proix et al., 2017), possibly supported by the alteration of whole-brain structural networks observed in epileptic patients (Besson et al., 2017). Our results further indicate that the network influence is strong enough not only to predict where a seizure will spread from known origin, but also in some cases identify this epileptogenic zone even if it is not directly discovered by the implantation only due to the effects it has on the connected observed regions (Fig. S4).

### 3.3. Reliance on the incomplete observations

Given that the method is trying the fill in incomplete data, it should be no surprise that there are limits of what can be inferred. Estimation of the onset times of hidden regions are only probabilistic and sometimes inaccurate, and the hidden epileptogenic zones can be discovered only when they are well connected to the observed regions. The straightforward way to reduce the uncertainty in the inference results is to observe more network nodes, however, the number of implanted electrodes is limited due to the clinical considerations. Other ways may be possible though. Even when the number of the observed nodes is fixed, it is possible that the uncertainty could be reduced by careful selection of the observed nodes before the implantation. The question of the system observability and the choice of the optimal sensor placement in complex networks is a problem well investigated in the field of control theory (Liu and Barabási, 2016; Aguirre et al., 2018). However, most results pertain to linear systems or nonlinear systems with polynomial or rational coefficients, and are thus not applicable to our system based on the threshold dynamics, especially considering that the trajectory through the high-dimensional state space cannot be *a priori* estimated. Furthermore, efficient analysis of large networks still poses a research problem.

Other way to increase the number of observed nodes would be to use more advanced technique to invert the SEEG signals to the space of brain regions, compared to here employed nearest-region mapping. Studies have demonstrated that source localization techniques may be beneficial not only in their traditional applications with non-invasive EEG and MEG recordings, but also with invasive recordings for interictal spike localization (Caune et al., 2014; Cam et al., 2017). Extending such approach to ictal recordings could lead to better exploitation of the data and thus to more precise and extensive seizure observations on the level of brain regions.

### 3.4. Assumptions of the method

The method introduced here rests on several assumptions regarding the nature of seizure dynamics. The guiding principle during the method development was simplicity over complexity with possibly irrelevant details; that was in order to build and analyze a transparent and robust method on which further, more refined versions can be based in the future. Some of the assumptions taken can be however questioned and we discuss them here.

#### Binary healthy/seizure state

Our method rests on the notion that the state of a brain region during a seizure can be clearly separated to either healthy or seizure state. That is an assumption shared with other network models of seizure propagation; variations on a dynamical system with a fixed point and a limit cycle representing the healthy and the seizure state respectively are commonly used in the network models (Goodfellow et al., 2016; Sinha et al., 2017; Jirsa et al., 2017). It is a useful conceptual simplification, but it a simplification nevertheless. While the electrographic recordings in human patients are commonly divided into interictal, preictal, ictal, and postictal periods, their boundaries or even their existence may not always be clear (Fisher et al., 2014). Multiple different electrographic patterns are observed in intracranial recordings at seizure onset (Perucca et al., 2014; Jiménez-Jiménez et al., 2015; Lagarde et al., 2016), which likely reflect different underlying dynamics of the neuronal tissue during seizure initiation and spread (Wang et al., 2017; Liou et al., 2020). Recent experimental studies paint a complex picture of spreading seizures with ictal core organized by fast traveling waves, ictal wavefront, and surrounding areas affected by strong feedforward inhibition (Schevon et al., 2012; Smith et al., 2016; Martinet et al., 2017; Liou et al., 2018). All of these can be considered abnormal states, yet all are distinct with different roles in seizure spread and with different contributions to the electrographic signals. As it is, the present inference method relies on the model simplicity obtained by having only two states. Nevertheless, introducing more complex dynamics while retaining the conceptual simplicity can be envisioned and should be pursued as the role of the different dynamical regimes in seizure organization and spread as well as their relation to the observable signals becomes clearer.

#### Normal distribution of the excitabilities

We set the prior distribution of the regional excitabilities to a normal distribution. This choice can be justified from two perspectives. First is that of information theory: the normal distribution has maximum entropy among the distributions with known mean and variance (Cover and Thomas, 2006) and thus in this sense it is the weakest assumption that can be made. Second, the regional excitability is an abstraction representing the cumulative effect of the underlying components that play a role in the ictogenesis. By virtue of the central limit theorem (Feller, 1968), if these are independent random variables, then their sum converges to the normal distribution, no matter what their original distribution is. It is however worth considering that localized structural abnormalities such as focal cortical dysplasias or brain tumors are among the common causes of epilepsies (Scheffer et al., 2017; Devinsky et al., 2018). While in such cases a clear distinction between the healthy and the affected tissue exists and a bimodal prior distribution might seem more appropriate, quantitative analysis of the intracranial signals indicates that also the structures outside of the lesion might have elevated epileptogenicity (Aubert et al., 2009; Sevy et al., 2014), therefore, the relation between the structural abnormalities and epileptogenicity is not as direct.

#### Unbiased implantation

The inference method introduced here implicitly assumes an unbiased implantation of the intracranial electrodes, i.e. that the observed regions are selected randomly. That is decidedly untrue, as the electrodes are implanted in the regions suspected of the seizure involvement based on the existing non-invasive data (Bartolomei et al., 2017; Parvizi and Kastner, 2018). Presumably it is this assumption that leads to the inferred widespread recruitment patterns such as shown on Fig. S8. Technically this bias can be implemented in the model by adding an appropriate constraint into the statistical model, however two problems remain. First, the implantation might not always perfectly correspond to the clinical hypothesis of involvement; some suspected regions might not be implanted due to the practical constraints of the electrode implantation such as the avoidance of blood vessels, safe distance between electrodes, or suitable entry angle through the skull. Second, and more fundamental, is the issue of validation. Any prior placed specifically on the hidden regions will primarily affect those hidden regions, and its effect on the observed regions will be only secondary and presumably minor. However, in the utilized leave-one-out framework, only the observed regions can be left out and thus only the predictions for the observed regions can be validated. In other words, current framework is not sufficient to properly evaluate the effects of the biased implantation assumption, and other approach would be needed.

#### Shared seizure dynamics

Focal seizures exhibit considerable variability between patients in their causes (Scheffer et al., 2017; Devinsky et al., 2018), electrographic onset patterns (Perucca et al., 2014; Lagarde et al., 2016), their duration (Jenssen et al., 2006), or underlying dynamic (Karoly et al., 2018). Even in individual patients, while more stereotypical (Kramer et al., 2010; Burns et al., 2014; Wagner et al., 2015), the seizures may still differ markedly in the extent of the recruitment or in the seizure duration (Jenssen et al., 2006; Martinet et al., 2017; Schroeder et al., 2019). In our model we explain this variability by the seizure-specific excitabilities, and, to a lesser degree, by patient-specific structural connectomes. It is possible that such approach is not sufficient to fully capture the variability of seizure dynamics, and that varying the excitation function *f*_***q***_ across the seizures and patients would lead to better results. Multiple options to do so are at hand, including the inference of the parameterization ***q*** on a single seizure or single patient level, or inferring the parameterization for specific seizure classes, either predefined or extracted from the data via appropriate unsupervised learning method.

### 3.5. Conclusions

Wide range of models of seizure dynamics exist today, ranging from detailed single-neuron models to network-based whole brain models (Wendling et al., 2016), with more recent studies attempting to link the models to patient-specific intracranial recordings via parameter inference on regional scale (Papadopoulou et al., 2017; Karoly et al., 2018) or at a whole-brain scale (Hashemi et al., 2020). In this work we introduce a method of the latter type. Here the seizure dynamics is extremely simplified, however, we do not discard the considerable complexity of seizure dynamics on micro- and meso-scale levels. We rather explore how much of the observed spatio-temporal organization can be explained by the simple principles encoded in our model that hides this complexity behind a single regional excitability parameter. In exchange for this simplification we obtain a model that can be reliably inverted and can exploit the intracranial electrographic data on the whole-brain scale not only for a single patient, but also for patient cohorts. The epilepsy models at different scales are mutually complementary, and it is by bridging the gap between the different levels that the patient-specific seizure dynamics on the whole-brain scale can be understood. We expect and hope that as the understanding of epileptic seizures on smaller scales progresses, its incorporation in the whole-brain models such as the one presented here will lead to the desired goal.

## 4. Materials and Methods

### 4.1. Patients and data acquisition

Fifty epileptic patients who underwent standard clinical evaluation for surgery candidates at La Timone hospital in Marseille were selected for the inclusion in this study. Details of the subjects are given in Supplementary Table S3. The evaluation included non-invasive T1-weighted imaging (MPRAGE sequence, either with repetition time = 1.9 s and echo time = 2.19 ms or repetition time = 2.3 s and echo time = 2.98 ms, voxel size 1.0 × 1.0 × 1.0 mm) and diffusion MRI images (DTI-MR sequence, either with angular gradient set of 64 directions, repetition time = 10.7 s, echo time = 95 ms, voxel size 1.95 × 1.95 × 2.0 mm, b-weighting of 1000 s mm^−2^, or with angular gradient set of 200 directions, repetition time = 3 s, echo time = 88 ms, voxel size 2.0 × 2.0 × 2.0 mm, b-weighting of 1800 s mm^−2^). The images were acquired on a Siemens Magnetom Verio 3T MR-scanner.

The invasive evaluation consisted of implantation of multiple depth electrodes, each containing 10 to 15 contacts 2 mm long and separated by 1.5 or 5 mm gaps. The SEEG signals were recorded by a 128 channel Deltamed system using at least 256 Hz sampling rate. The recordings were band-pass filtered between 0.16 and 97 Hz by a hardware filter. Only the seizures with duration longer than 30 seconds were included in the analysis. For some patients only these short seizures were recorded; their structural data were used in the study nevertheless. After the electrode implantation, a CT or T1-weighted scan of the patient’s brain was acquired to obtain the location of the implanted electrodes.

The patients signed an informed consent form according to the rules of local ethics committee (Comiteé de Protection des Personnes Sud-Meéditerranée I).

### 4.2. Structural model of the brain

The structural connectome was built with a reconstruction pipeline using generally available neuroimaging software. The current version of the pipeline evolved from a previously described version (Proix et al., 2016).

First, the command *recon-all* from FreeSurfer package (Fischl, 2012) in version v6.0.0 was used to reconstruct and parcellate the brain anatomy from T1-weighted images. For reasons of conformity with established implantation and interpretation practices at the epileptology department of AP-HM, we employed a custom brain parcellation in this study. This custom parcellation is based on the default FreeSurfer segmentation of the brain tissue (Fischl et al., 2002) with the Destrieux cortical parcellation (Destrieux et al., 2010), where some regions are merged together, some are split into multiple parts, and yet others are split and then merged with existing ones. The full list of the performed operations is given in Supplementary Table S1. The resulting parcellation contains 162 regions with 72 cortical and 9 subcortical regions per each hemisphere (Supplementary Table S2).

Next, the T1-weighted images were coregistered with the diffusion weighted images by the linear registration tool *flirt* (Jenkinson et al., 2002) from FSL package in version 6.0 using the mutual information cost function with 12 degrees of freedom. The MRtrix package in version 0.3.15 was then used for the tractography. The fibre orientation distributions were estimated from DWI using spherical deconvolution (Tournier et al., 2007) by the *dwi2fod* tool with the response function estimated by the *dwi2response* tool using the *tournier* algorithm (Tournier et al., 2013). Next we used the *tckgen* tool, employing the probabilistic tractography algorithm iFOD2 (Tournier et al., 2010), to generate 15 millions fiber tracts. The connectome matrix was then built by the *tck2connectome* tool.

Each element *ŵ* _*ij*_ of the generated connectome matrix *Ŵ* represents the number of fibers from region *j* to region *i*. The elements of the matrix were then scaled by the volumes of the target regions, 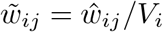, where *V*_*i*_ is the volume of *i*-th region. This way, the elements 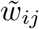 represent the density of the fibers projecting from region *j* in region *i*. Afterwards, a correction for hippocampus connections was introduced: our custom brain atlas has the hippocampus split into two brain regions - anterior and posterior hippocampus. To correct for the presumably strong gray matter connections between the two parts, which are not discovered by the white matter tractography, the connectome matrix elements corresponding to the connection between the anterior and posterior hippocampus are increased by the value of 98 percentile of the connectome weights. Finally, the connectome is normalized so that the maximal sum of ingoing projections is equal to one: 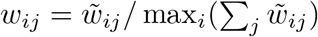

The electrodes contacts were localized using GARDEL software (Medina Villalon et al., 2018), available at https://meg.univ-amu.fr/wiki/GARDEL:presentation. Using the CT scan, the software automatically detects all SEEG electrodes and localizes the belonging contacts. These coordinates were then projected back to the reference frame of T1-weighted images using the transformation obtained by the linear registration tool *flirt*.

### 4.3. Resection masks and surgery outcome

For the patients who underwent resective surgery, the extent of surgical resection with respect to the anatomical parcellation according to the in-house custom atlas was defined using the EpiTools software suite (Medina Villalon et al., 2018). In brief, patient’s specific 3 D m aps w ith SEEG electrodes were created and all the contacts were assigned to the anatomical regions of the atlas projected in the pre-operative MRI space. The co-registration of the post-implantation CT scan with electrodes with the post-operative MRI scan, and that of the post- and pre-operative MRI scans were then performed. The resected contacts were identified and assigned to the respective regions as defined by the p re-operative MRI. The regions resected b ut not e xplored by depth e lectrodes were then identified on the pre-operative/post-operative MRI co-registered images manually (SMV, JS). The completeness of resection of each respective region was estimated visually by two trained clinicians (FB, JS) and expressed in percentages. For the purpose of comparison with model predictions, only the regions with the resection extent above 50% (non-inclusive) were considered.

The surgery outcome was classified using the Engel score, w here class I corresponds to patients free of disabling seizures, class II to patients with rare disabling seizures, class III to worthwhile improvement, and class IV to no worthwhile improvement.

### 4.4. Onset time detection

The onset times were detected on the SEEG recordings in bipolar representation. For each bipolar channel, the onset time was detected by the following sequence of steps: First, the log-power in two frequency bands (*LP*_low_(*t*) from 1 to 12.4 Hz; *LP*_high_(*t*) from 12.4 to 100 Hz) was calculated by computing the time-frequency representation using the multitaper method, summing over the given frequency band. Next, the log-power time series were normalized to preictal baseline, determined as the mean log-power in the 60 seconds preceding the seizure onset as marked by a clinician:

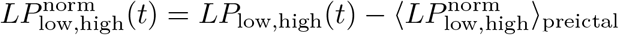

Then the binary seizure mask was created:

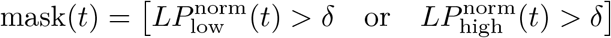

where [*P*] is the Iverson bracket,

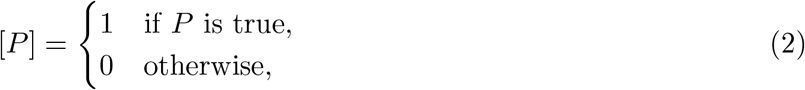

and *δ* = log(5) is the prescribed threshold, corresponding to five-fold increase (or decrease) of power in a given band. Next, the mask was cleaned up to remove short intermittent intervals of seizure and healthy states. The mask was first smoothened by convolution with a rectangular window of duration 20 seconds and then binarized again with threshold 0.5. Finally, intervals of seizure state shorter than 20 seconds were removed. If there was no remaining seizure interval, then the channel was declared as non-seizing. Otherwise it was declared as seizing with the channel onset time equal to *t*_*ch*_ = min{*t* | mask^clean^(*t*) = 1}.

### 4.5. Mapping of channel observations to brain regions

The channel onset times were mapped onto the brain regions using the physical distance of the electrode contacts and the voxels belonging to the regions. For each bipolar channel, we took the position of the midpoint between the two electrode contacts, and we computed the distance to the brain regions. If the two nearest regions were in a similar distance from the midpoint (i.e. satisfying *d*_2_*/*(*d*_1_ + 0.5mm) < 2, where *d*_1_ and *d*_2_ are the distances of the nearest and second nearest region respectively), we did not assign the observation to any region, otherwise we assigned it to the closest region.

After the assignment is done, none, one, or multiple observations may be assigned to a single region. If there was no observation, the region was considered hidden. If there was one, region observation was set to the assigned channel observation, and if there were multiple observations assigned, the region observation was set as the median of the assigned observations (with lower interpolation if there was an even number of observations). To facilitate the median calculation, the non-seizing regions were considered to be seizing at *t* = ∞.

If there were no seizing regions after the mapping, we excluded the seizure from further analysis. Otherwise, the onset times were shifted so that the first region onset time was at *t* = 30 s, and the regions with onset time greater than *t*_lim_ = 90 s were set as non-seizing.

### 4.6. Dynamical model of seizure propagation

#### The model

At the core of the method is the following model of seizure propagation in a weighted network. For a network with *n* regions, the model reads

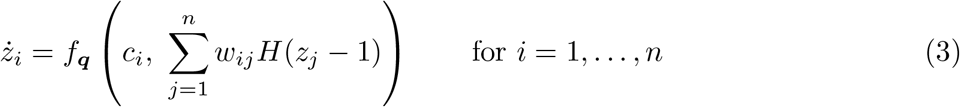

Here *z*_*i*_ is the slow variable of region *i*. Parameter *c*_*i*_ is the node excitability, and *W* = (*w*_*ij*_) is the connectivity matrix, normalized so that max_*i*_ ∑_*j*_ *w*_*ij*_ = 1 as described in Sec. 4.2. The function *f*_***q***_ : ℝ × [0; 1] → ℝ^+^ is the *excitation* function, parameterized by the parameter vector ***q***. Due to the scaling of the connectome, the second argument is guaranteed to fall in the interval [0, 1]. We require that the function *f*_***q***_ is positive and increasing w.r.t. its first parameter. The positivity requirement guarantees that the slow variable can only increase and thus all regions are pushed only closer to the seizure state. The increasingness in *c* is the only requirement that breaks the symmetry in *c*, and it assures that the larger values of *c* can be interpreted as more excitable. We furthermore require that *f*_***q***_(*c, y*) is onto ℝ^+^ for any *y*, that is to assure the existence of a solution to the inverse problems (see SI Text). Finally, *H* is the Heaviside step function, representing the switch from the healthy to the seizure state when the slow variable crosses the threshold *z* = 1. The onset time of a region *i* is defined as *t*_*i*_ = min{*t* | *z*_*i*_(*t*) ≥ 1}. The system is completed by the initial conditions *z*_*i*_(0) = 0. For a known vector of excitabilities ***c***, parameter vector ***q***, and a connectome matrix *W*, the model uniquely defines the vector of onset times ***t***. We use the following shorthand for this mapping:

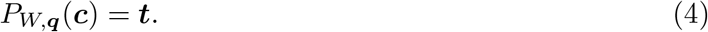

Because the function *f*_***q***_ is positive, the model implies that every region will start to seize at some finite time. In reality, this is not the case, and to account for that in the statistical model we will introduce a time limit of a seizure, and consider all regions with onset time larger than the limit to be non-seizing.

#### Parameterization of the excitation function f_**q**_

In general, the excitation function *f*_***q***_(*c, y*) can be parameterized in any fashion, provided that it guarantees its positivity and its increasingness in the first argument. In this work we opted for a simple approach of a bilinear function followed by an exponentiation. Given the real values of the bilinear function *q*_*aa*_, *q*_*ab*_, *q*_*ba*_, *q*_*bb*_ in the interpolation points [*c*_*a*_, *y*_*a*_], [*c*_*a*_, *y*_*b*_], [*c*_*b*_, *y*_*a*_], [*c*_*b*_, *y*_*b*_] with *c*_*a*_ = −1, *c*_*b*_ = 1, *y*_*a*_ = 0, *y*_*b*_ = 1, the function reads

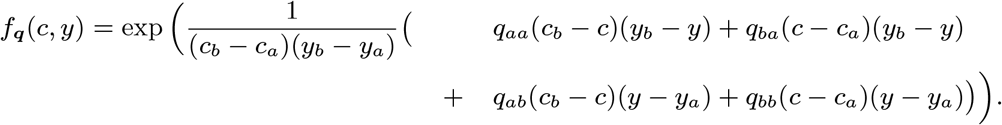

In order to assure that *f*_***q***_ increases in *c*, we declare 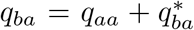 and 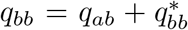, and we parameterize the function with the parameter vector 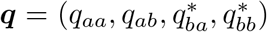 where *q*_*aa*_, *q*_*ab*_ ∈ ℝ and 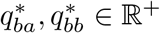.

### 4.7. Statistical model and the inference

Two statistical models are used in the study. The hierarchical model for multi-seizure inference (Box 1) is used for the learning of the hyperparameters. It takes the data from multiple seizures and infers the parameters ***q*** of the excitation function *f*_***q***_. The low-level parameters - excitabilities ***c***_*k*_ for all seizures *k* - are inferred as well, but due to the design of the leave-one-out validation scheme are not used in this study. The prior for the hyperparameters and its variance *σ*_*q*_ specifically was selected as noninformative based on the observation that the parameters sampled from this prior distribution can produce wide range of behavior, including synchronized cascades of region onsets or nonsynchronous onsets in a model with no network effects.

The model for single-seizure inference (Box 2) is the simplification of the multi-seizure model, where the hyperparameters are fixed to given values, and the parameters of only one seizure are inferred. In this study, this model is used for the leave-one-out validation and the validation against the resection, where the hyperparameters are fixed to values learned by the multi-seizure model on training data.

The parameter inference with both multi- and single-seizure models was performed using the No-U-Turn Sampler (Hoffman and Gelman, 2014), a self-tuning variant of Hamiltonian Monte Carlo method (Neal, 2011; Betancourt, 2017) that eliminates the need to set the algorithm hyperparameters. The implementation in Stan software was used (Carpenter et al., 2017).

For the multi-seizure inference, four independent MCMC chains were run each with random initiation in search space, and 500 steps in the warm-up phase and 500 generated samples. For the single-seizure inference, two chains were run, again with 500 steps in the warm-up phase and 500 generated samples.

#### Box 1.

Statistical model for multi-seizure inference

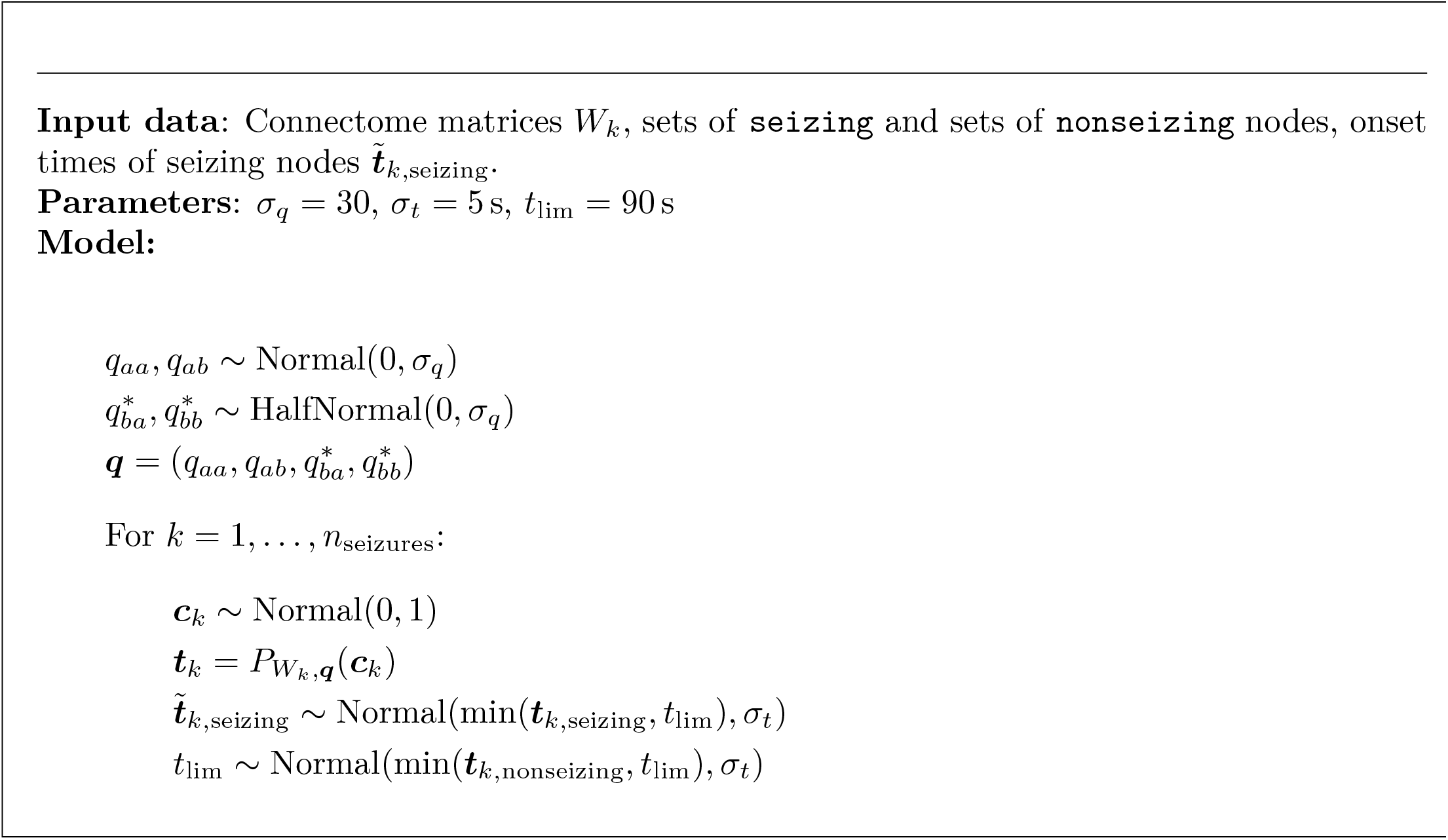

#### Box 2.

Statistical model for single-seizure inference

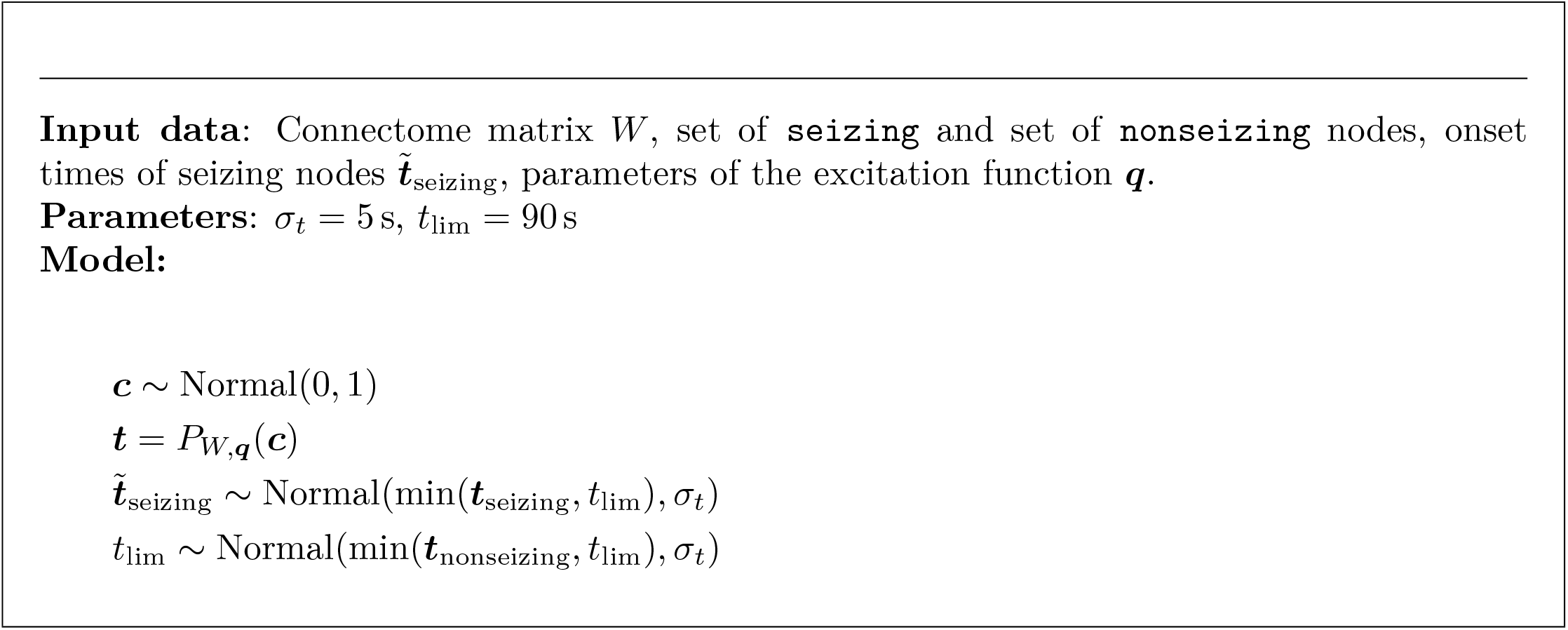

### 4.8. Validation on real data

The leave-one-out cross-validation and the validation against the resection masks consisted of the following steps: First, we detected the onset times in all recorded seizures longer than 30 seconds, and excluded the seizures where no seizing region was detected. Next, the cohort of 50 subjects was divided in two folds (each with 25 subjects) and for both of these two folds the multi-seizure model was fitted independently of the other. At most two seizures from a single subject were included in the training data to prevent unproportional influence of any subject on the group results; if more than two seizures were available for one subject, the two seizures were selected randomly. In each fold the posterior distribution of the hyperparameters ***q*** was obtained, and its mean was taken as a point estimate of the hyperparameters used in the subsequent steps.

With the inferred hyperparameters the leave-one-out validation was performed. For each fold, all seizures were repeatedly fitted with a single-seizure model, each time using the seizure data set with observation of one region excluded. The inferred states and onset times of the excluded regions were then compared to the left-out observations and evaluated using the measures described below. The hyperparameter values used for the single-seizure fitting in one fold were those obtained from the other fold, thus preventing reusing the same data for the model training and validation. To get the excitabilities used in the validation against the resection and surgery outcome, all seizures from both folds were fitted using the single-seizure model once more, this time with no observations left out. Again, the hyperparameter values were those obtained from the other fold. The entire computational pipeline was constructed and run using the Snakemake workflow manager (Köster and Rahmann, 2012).

### 4.9. Evaluation measures

#### State and onset prediction accuracy

We define the *state prediction accuracy* for a region *i* as the predictive probability that the region state is equal to the ground truth state conditioned on the data. We approximate this quantity using the posterior onset time samples, i.e.

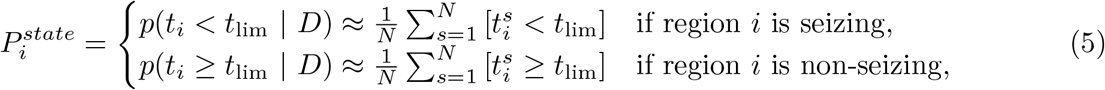

where *t*_*i*_ is the onset time of region *i, D* represents the onset times data (either from all observed regions for synthetic data validation, or with the region *i* excluded for leave-one-out validation with empirical data 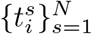 are the posterior samples of the onset time of region *i* with the superscript *s* indexing over *N* samples, the constant *t*_lim_ is the artificial limit of a seizure, and [·] is the Iverson bracket (2). The motivation for this definition is the association of the regions that start to seize late (after time *t*_lim_) with non-seizing regions. The ground truth state is either the known simulated state of a hidden region in case of the synthetic data validation, or the state of a left-out observed region in case of the leave-one-out cross-validation on real data.

We define the *onset prediction accuracy* for a seizing region *i* as the predictive probability that the onset time *t*_*i*_ is sufficiently close to the ground truth conditioned on the data,

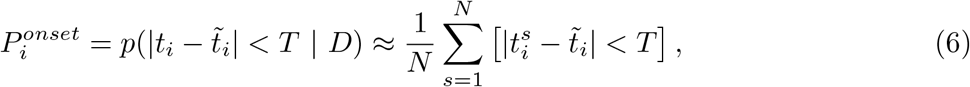

where 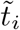 is the ground truth, which is either the known simulated onset time in case of the synthetic data validation, or the left-out onset time in case of the leave-one-out validation on real data. The constant *T* determines the temporal resolution of the measure, in this work we use *T* = 5 s. To avoid the border effect affecting the seizing regions with onset time close to *t*_lim_, we evaluate the onset prediction accuracy only for the seizing regions with 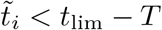.

#### Estimation methods

To provide a point of comparison for the state and onset prediction accuracies obtained with the inference method, we introduce two simpler estimates of the region onset times. The calculation of the state and prediction accuracies in (5) and (6) for region *i* uses the set of posterior samples of the onset time 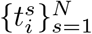 In the (unweighted) estimate we replace this set with the observed onset times of all other observed regions in the same seizure 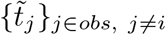 where *obs* denotes the set of observed regions. For the purpose of this computation we set the onset time of the observed non-seizing nodes to infinity. The state and onset predicted accuracies are thus calculated as

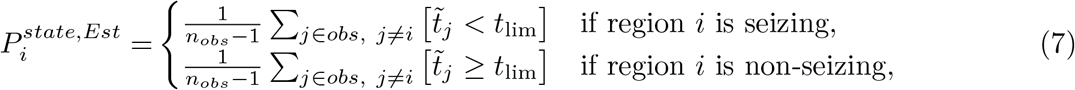

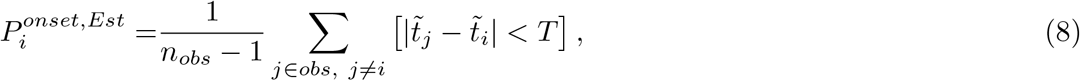

In the weighted estimate we replace the set of posterior samples for region *i* in (5) and (6) with a theoretical set of the onset times of the other observed regions *j* in the same seizure, where every onset time is repeated proportionally to the weight of the connection between regions *i* and *j*. In practice we do not build this set, we simply replace the averages in (7) and (8) with their weighted counterparts,

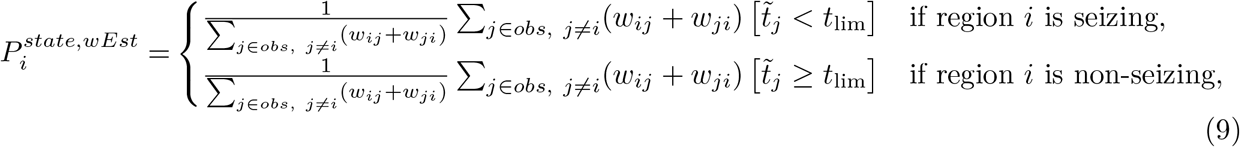

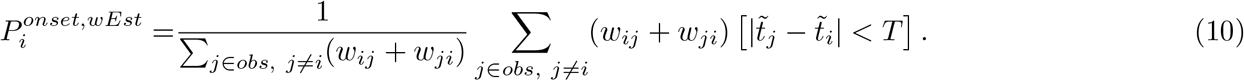

#### Precision and recall

To compare how the predicted epileptogenicity correspond to the ground truth epileptogenicity (in case of synthetic data) or to resected regions (in case of real data) we use the measures of precision and recall. In this work, the predictive variable is the posterior probability thata region *i* is highly excitable 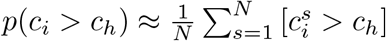 where *c*_*h*_ = 2 is the threshold of high excitability. Given a threshold for the predictive variable we obtain a binary vector of predictions, which is compared with the binary vector of relevant elements. These are either the epileptogenic regions (for synthetic data) or resected regions (for real data). The comparison gives the number of true positives (TP; predicted and relevant regions), true negatives (TN; not predicted and not relevant regions), false positives (FP; predicted and not relevant regions), and false negatives (FN; not predicted and relevant regions). The *precision* is defined as 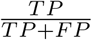 and *recall* as 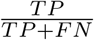. These measures are well-suited for imbalanced data sets, where number of true negatives outweighs the other categories, as the number of true negatives does not enter in the calculation of the precision nor of the recall. Indeed, epileptogenic zone prediction is an example of such imbalanced data set, as the number of predicted and relevant regions is generally small relative to the number of brain regions.

## Data Availability

The patient data sets cannot be made publicly available due to the data protection concerns,
but the anonymized data can be shared by authors upon reasonable request.

## Author contributions

VS and VJ designed the study. HF, JS, SMV, MG, and FB collected and organized the data. SMV, JS, and FB prepared the resection masks. VS developed the method and performed the computational analysis. VS, AV, MH, MW, and VJ analyzed the results. VS drafted the manuscript. All authors edited the manuscript.

## Acknowledgements

The authors wish to acknowledge the financial support of the following agencies: the French National Research Agency (ANR) as part of the second “Investissements d’Avenir” program (ANR-17-RHUS-0004, EPINOV), European Union’s Horizon 2020 Framework Programme for Research and Innovation under the Specific Grant Agreement No. 785907 and 945539 (Human Brain Project SGA2 and SGA3), the Fondation pour la Recherche Medicale (DIC20161236442), VirtualBrainCloud (grant number 826421), Swiss National Supercomputing Centre (CSCS) under project ID ich001, and the SATT Sud-Est (827-SA-16-UAM) for providing funding for this research project.

## Data and code availability

The patient data sets cannot be made publicly available due to the data protection concerns, but the anonymized data can be shared by authors upon reasonable request. The code will be made available upon publication.

## Supplementary information

### Note on parameter identifiability

Investigation of the properties of the dynamical model can shed light on the identifiability of the parameters of the single-seizure statistical model (Box. 2) in the limit case of no observation noise *σ*_*t*_ = 0. Recall that the dynamical model reads

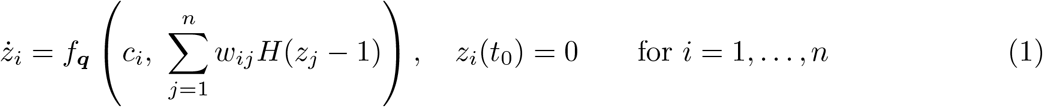

with *t*_0_ = 0, and that the onset time of a region *i* is defined as *t*_*i*_ = min {*t* | *z*_*i*_(*t*) ≥ 1}. Recall also that we use the shorthand *P*_*W*,***q***_(***c***) = ***t*** for the mapping between the excitabilities ***c*** = (*c*_1_, …, *c*_*n*_) and onset times ***t*** = (*t*_1_, …, *t*_*n*_) defined by the dynamical model (1) with the connectome matrix *W* and parameterization ***q***.

**Theorem 1**. *Let f*_***q***_(*c, y*) : ℝ *×* [0, 1] → ℝ^+^ *be a continuous function, strictly increasing in c and onto* ℝ^+^ *for any y. Then P*_*W*,***q***_ : ℝ^*n*^ → (ℝ^+^)^*n*^ *is one-to-one and onto*.

*Proof*. Since *z*_*i*_(*t*_0_) = 0 and *z*_*i*_(*t*) is increasing in *t* due to the positive RHS of (1), it follows from the definition that *t*_*i*_ ∈ ℝ^+^ for all *i* = 1, …, *n*. One thus has to show that for every ***t*** *∈* (ℝ^+^)^*n*^ there exists one and only one ***c*** *∈* ℝ^*n*^ that satisfies *P*_*W*,***q***_(***c***) = ***t***.

Assume that there is a ***t*** = (*t*_1_, *t*_2_, …, *t*_*n*_) *∈* (ℝ^+^)^*n*^, and assume, without loss of generality, that it is ordered so that 0 *< t*_1_ ≤ *t*_2_ ≤ … ≤ *t*_*n*_. If not, one can simply reorder the indices of the regions. Since the RHS in (1) is constant for *t ∈* [*t*_*j*−1_; *t*_*j*_) for any *j* = 1, …, *n*, one can write

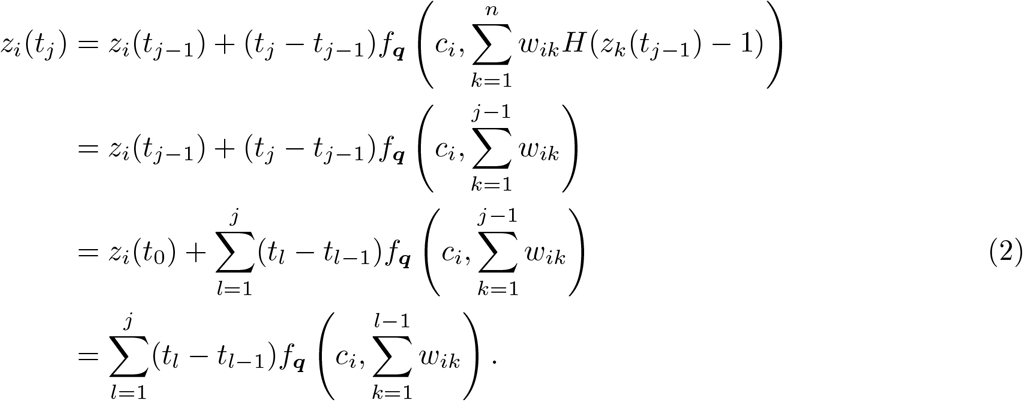

The second equality follows from the definition of *t*_*k*_ implying that *z*_*k*_(*t*) *<* 1 for *t < t*_*k*_ and from the ordering of ***t***. By setting *i* = *j* we obtain

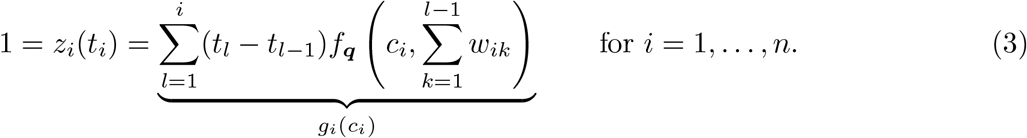

Since (*t*_*l*_ − *t*_*l −* 1_) are non-negative (with at least *t*_1_ − *t*_0_ being positive) and *f*_***q***_ is strictly increasing in *c* and onto ℝ^+^, it follows that *g*_*i*_ is also strictly increasing and onto ℝ^+^, therefore (3) has a unique solution for all *i* = 1, …, *n*.

This theorem helps to make a clear link between the observations and the model parameters.

In an observation of a spreading seizure in a network any region *i* can be either

- observed seizing, in which case it has a known onset time *t*_*i*_;
- or observed non-seizing, which we represent as having an onset time *t*_*i*_ ≥ *t*_lim_;
- or hidden.

If we reorder the regions in this order, any particular observation of a seizure can be represented as a set *T* in the space of observations (ℝ ^+^)^*n*^,

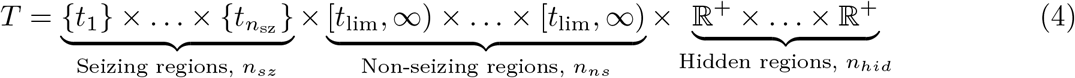

This is an (*n*_*ns*_ + *n*_*hid*_)-dimensional manifold, and if *n*_*ns*_ > 0, then it has an (*n*_*ns*_ + *n*_*hid*_ 1)-dimensional boundary representing the limit cases of one non-seizing region having the onset time*t*_lim_. And since *P*_*W*,***q***_ is one-to-one and onto, 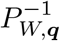 exists, and 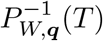 is a manifold of the same dimensionality in the space of excitability parameters ℝ^*n*^. This parameter manifold represents all possible solutions to the inverse problem for a particular observation *T*. The probability of the solutions on the manifold is then determined by the prior distribution on the parameters. Fig. S1 illustrates this in a three-node network.

#### Validation on synthetic data

To instill confidence in the inference process and to establish what can be expected from the inference from the incomplete seizure observations, we first performed extensive validation of the method on synthetic data, i.e. on data generated by the same dynamical model which was then used for the inference. For three different functions *f*_***q***_ (with no coupling, weak coupling, or strong coupling; Fig. S2) we generated synthetic seizures by randomly sampling the excitabilities from standard normal distribution and calculating the region onset times using the dynamical model. If there were no seizing regions in the simulated seizure (i.e. no regions with onset time *t*_*i*_ *< t*_lim_ with *t*_lim_ = 90 s), a new seizure with new excitability parameters was generated. The parameters ***q*** of the excitation functions were selected by hand, loosely guided by the principle of preserving some key statistics of generated seizures (Fig. S2B). The exception was the strong coupling function, which was in fact the result of the inference performed on real data; the results for strong coupling thus provide an estimate of what can be expected in real data. We then ran the multi-seizure and single-seizure inference on these synthetic data, assuming that only portion of the regions (21, 54, or 108 out of 162) were observed, and we evaluated how well were the original parameters recovered. For every seizure, first one observed region was selected from those seizing, and then the other observed regions were selected randomly from all regions; this was to assure that there was at least one region seizing among those observed.

To test the recovery of the hyperparameters ***q***, we ran the multi-seizure model on batches of twelve synthetic seizures. Four independent MCMC chains were run for each batch; out of total of 36 chains one chain was stuck and was excluded from further analysis. The convergence was evaluated using the split-chain reduction factors 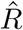 and number of effective samples *N*_*eff*_ (Gelman *et al*., 2013). All but one scenarios converged satisfactory with 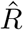 below 1.1 and with *N*_*eff*_ above 30 for all hyperparameters. The exception was the scenario with the strong coupling function with 108 observed regions 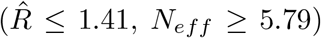; these results are nevertheless shown as well. adopt the evaluation framework of Betancourt (2018) and present the values of posterior z-score and posterior shrinkage. For a parameter *θ*_*i*_, the posterior z-score quantifies how far is the posterior from the ground truth,

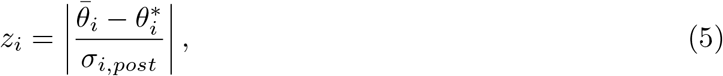

**Figure S1:**
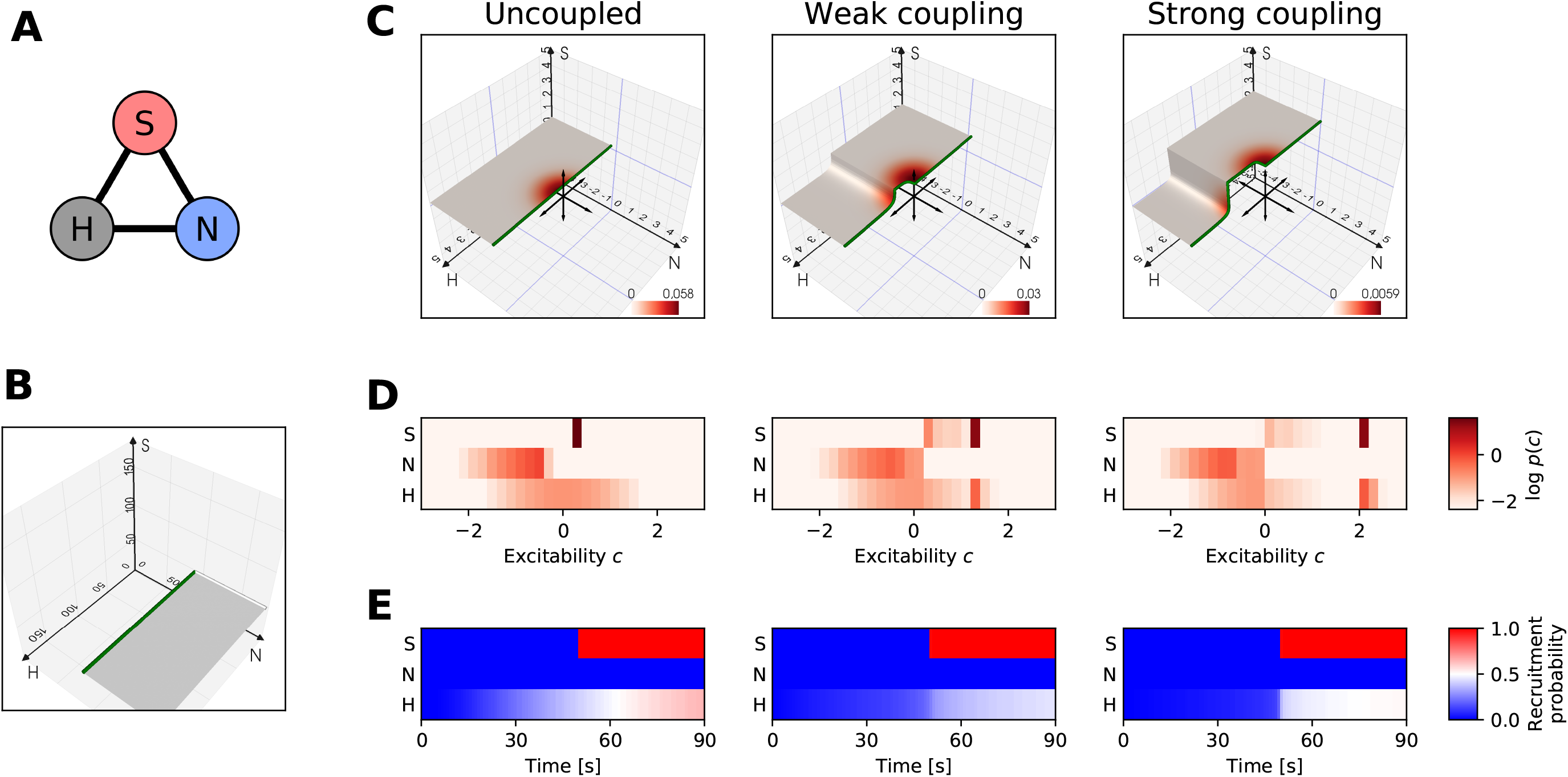
Observation and parameter manifolds in a three-node network. (A) Three node network with one hidden, one observed non-seizing, and one observed seizing region (with *t*_*S*_ = 50 s), marked with H, N, and S respectively. The network is fully and symmetrically connected with connection strengths *w*_*ij*_ = 0.1. (B) The observation manifold in the onset time space (ℝ^+^)^3^. The green line marks the limit case of *t*_*N*_ = *t*_lim_. (C) The parameter manifolds in the parameter space ℝ^3^ for three excitation functions *f*_***q***_ representing no, weak, and strong coupling (see Fig. S2). In general, the dimensionality of the manifolds is determined by the number of seizing, non-seizing, and hidden regions in the network, while the shape of the manifold is determined by the connectivity *W*, excitation function *f*_***q***_, and the onset times of seizing regions. The coloring represents the prior distribution of the excitability parameters, which is a standard normal distribution *N* (**0, *I***). (D) Marginal posterior probabilities of the excitability parameters of the three nodes, obtained by integrating the prior over the manifold. (E) Recruitment probabilities of the three regions. With no observation noise, the only differences are in the hidden region H, and the observations of the other two regions are reproduced exactly. This figure illustrates how the stronger coupling leads to more complicated posterior geometry (C), possibly resulting in a multimodality in the distribution of the excitability parameters (D). In this case, in addition to the dominant mode representing the highly excitable observed region S, the strong coupling introduces also a secondary mode representing the highly excitable hidden region H and less excitable observed region S. In other words, the coupling allows for a discovery of a hidden epileptogenic zone.

**Figure S2:**
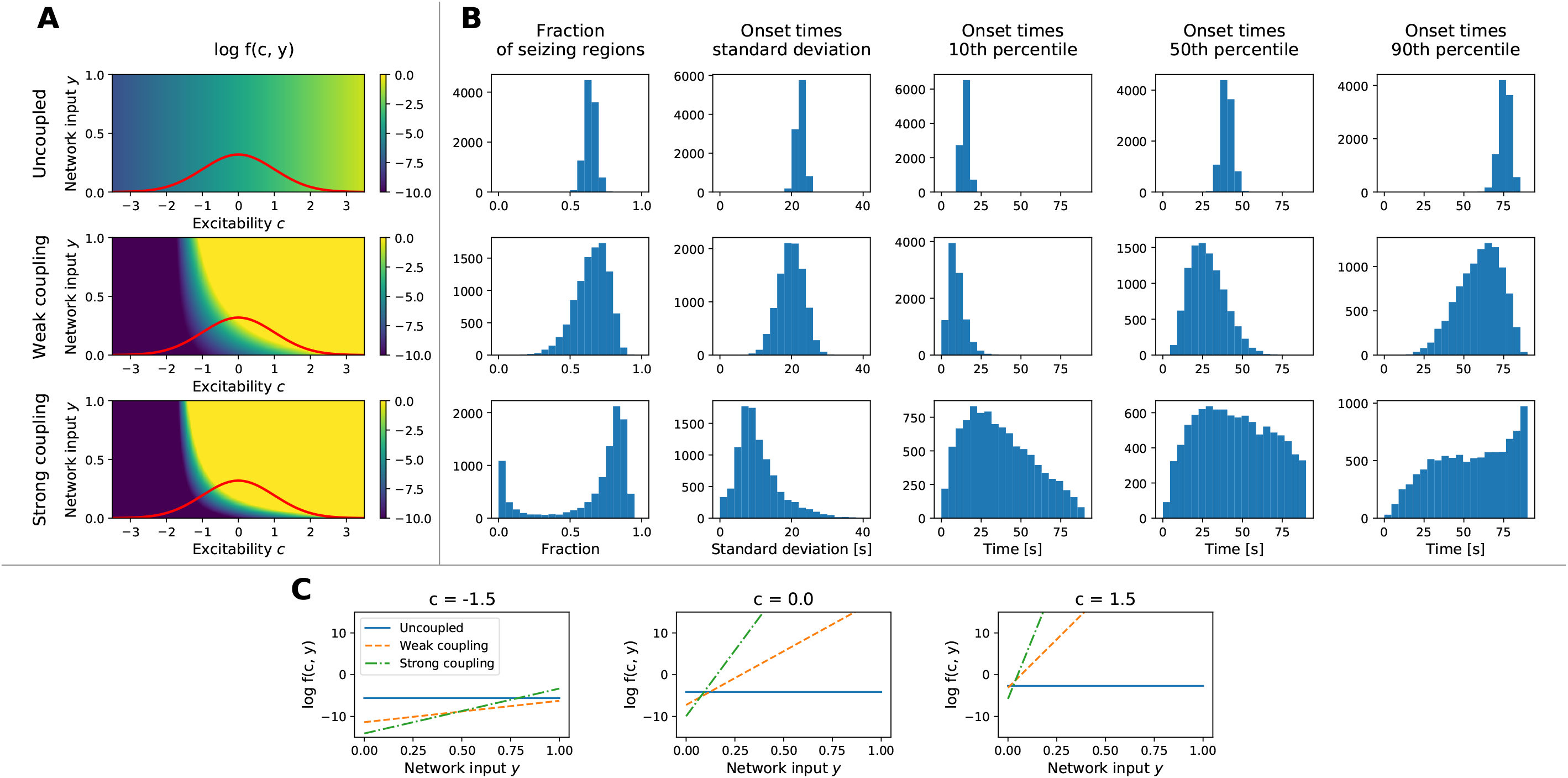
Three excitation functions used for the synthetic data validation. (A) Plots of the functions *f*_***q***_ (*c, y*) named “Uncoupled” with ***q*** = (5.12, 5.12, 1.95, 1.95), “Weak coupling” with ***q*** = (10.0, 2.0, 5.5, 33.0), and “Strong coupling” with ***q*** = (12.70, 15.48, 5.53, 75.21). The colormap is clipped to show the range of values relevant for the seizure propagation on timescales of seconds to tens of seconds. The red line shows the prior distribution on the region excitabilities *c*, i.e. the standard normal function. (B) Statistical quantification of the generated seizures. For each function (in rows) 10000 seizures were generated by taking a random patient connectome and randomly sampling excitabilities using the standard normal distribution. Columns show the histograms of properties of the generated seizures: fraction of seizing regions, standard deviation of the onset times of seizing regions, and percentiles of the onset times of seizing regions. Vertical axis always represents the count of seizures. (C) Dependency of the excitation functions on the network input for different values of excitability. The stronger or weaker relation motivates our notation of Uncoupled, Weak and Strong coupling for the different functions.

where 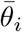 is the mean of the posterior distribution, 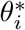 is the ground truth, and *σ*_*i,post*_ is the standard deviation of the posterior distribution. The posterior shrinkage quantifies how much is the uncertainty in the prior distribution reduced,

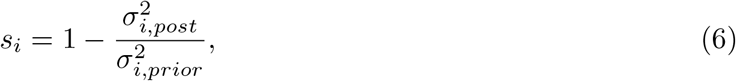

where *σ*_*i,prior*_ is the standard deviation of the prior distribution. Fig. S3A-C shows the results. Most data points are located in the lower right corner, indicating good recovery of the parameters. Some overfitting however can be seen for the uncoupled model; considering that the shrinkage values are very close to one, the distance is still small in the absolute numbers. This overfitting may be caused by the fact that the data were generated without any observation noise assumed in the statistical model, leading to more confident estimations.

To test the recovery of the low-level excitability parameters we ran the single-seizure model on synthetic seizures, assuming the knowledge of the hyperparameters ***q*** used for their generation. We have again used the same three sets of the hyperparameters and for each of them we generated 96 seizures, again sampling the excitability parameters randomly from a standard normal distribution, and using the connectomes from the patient cohort. One chain out of total of 1728 was stuck and was excluded. The vast majority of the inference runs converged well, with 99.84% regional excitability parameters with 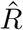 below 1.1 and *N*_*eff*_ above 30. We evaluated the relation between the ground truth and the mean of the posterior distribution, separately for the observed (Fig. S3D) and hidden nodes (Fig. S3E). For the observed nodes, best values are obtained in the uncoupled network, and the goodness of fit is reduced with stronger coupling; the effect of added observed nodes is minimal. For the hidden nodes the relation between the posterior mean and ground truth is in general much weaker. When the network is entirely uncoupled, nothing can be inferred about the hidden nodes. With stronger coupling some information can be extracted, with positive effect of added observed nodes. Interestingly, too strong coupling decreases the goodness of fit. In such scenarios the network acts homogeneously, and it is difficult to infer which node is the driver and which is the follower. Fig. S3F,G shows how well can be the seizing/non-seizing state and the onset times of hidden nodes predicted. Here the relation is clear: stronger coupling leads to better predictions, with small benefits of adding observed nodes.

#### Discovery of epileptogenic zones in synthetic seizures

Before applying the method to discover the epileptogenic zone in the patient recordings, we tested how well can the epileptogenic zones be discovered in the synthetic data. These were generated by the trained model using patients’ structural connectivities and electrode implantations. In the model, we equated the epileptogenic zone with the region with high excitability *c > c*_*h*_ for the threshold *c*_*h*_ = 2. In order to establish how well can be the epileptogenic zone discovered based on its location relative to the implanted sensors, we analyzed three scenarios: observed epileptogenic zone, hidden epileptogenic zone with random location among the non-observed regions, and a *near miss* scenario, that is a hidden epileptogenic zone that is well connected to at least three observed regions (Fig. S4A). We analyzed how well were the epileptogenic zones discovered with the precision-recall curve using the inferred probability of high excitability (Fig. S4B). The observed epileptogenic zones were well identified, while the identification of the hidden epileptogenic zone was unsurprisingly worse. However, the location matters considerably. In the random scenario, the precision and recall for the threshold *p*_*t*_ = 0.05 is 4% and 9% respectively. In comparison, the precision and recall in the near miss scenario is at 11% and 33%, meaning that a third of the hidden epileptogenic zones were discovered with above one in ten chance of being correct. While such numbers are far from being sufficient to serve as a sole basis for any surgical decision, it might be enough to point to previously unexpected cause for which supporting evidence may (or may not) be found by other methods.

**Figure S3:**
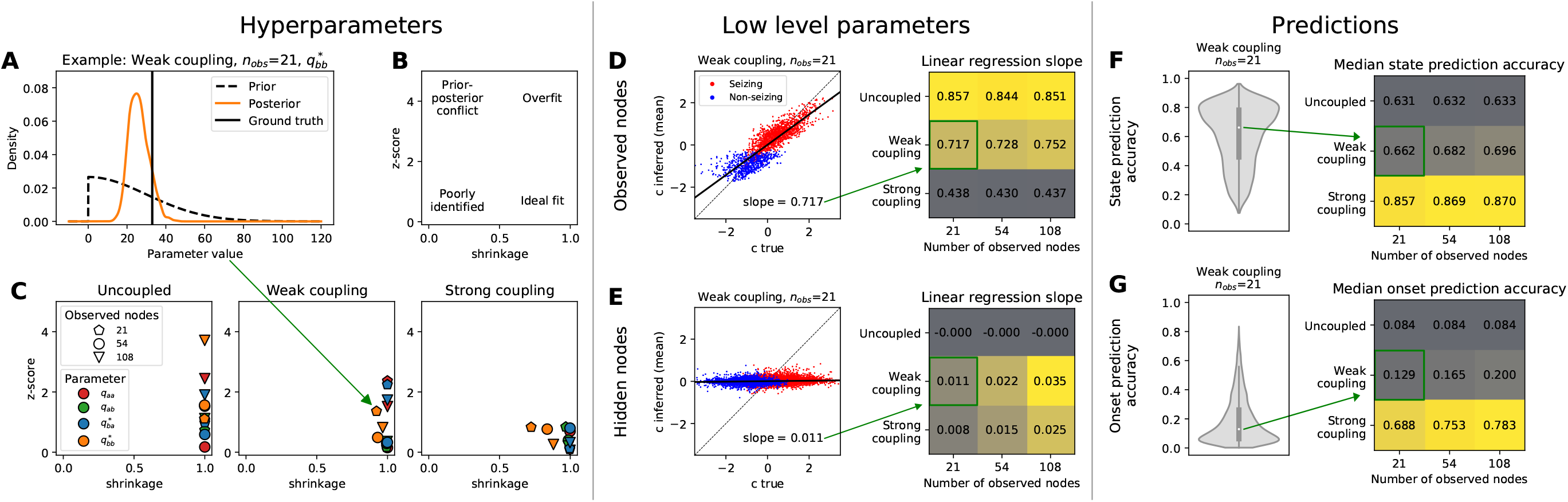
Validation on synthetic data. (A) Example of a hyperparameter inference. The posterior is more concentrated around the ground truth compared to the prior. (B) Interpretation of the shrinkage - z-score plot. Lower right corner constitutes the ideal fit. (C) Shrinkage - z-score plots for the three excitation functions, three numbers of observed nodes out of total of 162, and all four hyperparameters. (D,E) Goodness of fit for the low-level parameters. Left panels show the relation between the ground truth and the posterior mean of the excitability in one simulated scenario with weak coupling and 21 observed nodes. Each dot represent one brain region, colored based on whether it seizes or not. The dashed line represents a perfect fit and the solid line shows the linear regression fit. Right panels show the linear regression slope for all scenarios. Note that the color code is different between D and E. (F) Distribution of state prediction accuracy (i.e. of the probabilities that a seizing/non-seizing state of a hidden region is correctly inferred) for one scenario (left), and its median for all scenarios (right). (G) Distribution of onset prediction accuracy (i.e. of the probabilities that a onset time for a hidden seizing region is correctly inferred) for one scenario (left), and its median for all scenarios (right). The violin plots in F and G show a kernel density estimate of the probability density of a given variable across all regions and across all seizures for the given scenario. The inner boxplots show the median (white dot), interquartile range (IQR, gray bar) and adjacent values (upper/lower quartile +/−1.5 IQR, gray line).

**Figure S4:**
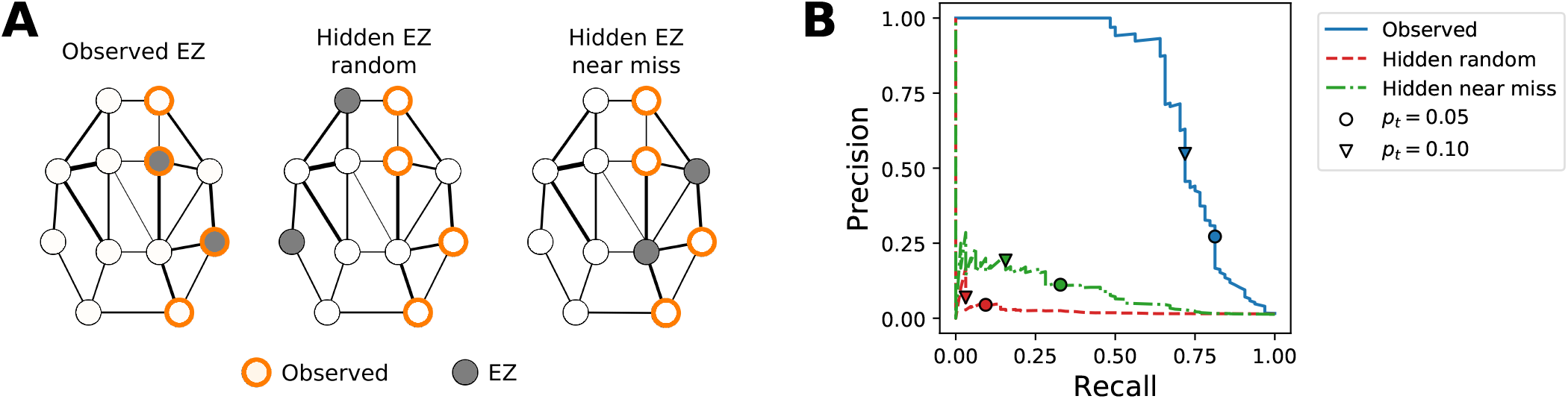
Discovery of the observed and hidden epileptogenic zones (EZ) in synthetic seizures. (A) Three scenarios of EZ location. In all three scenarios, two EZs (with the excitabilities drawn from the standard normal distribution bottom-truncated at *c* = *c*_*h*_) are placed in the network, and the rest of the nodes has excitabilities drawn from the standard normal distribution top-truncated at *c* = *c*_*h*_. In the observed EZ scenario, the EZs are placed randomly among the observed nodes. In the random hidden EZ scenario, the EZs are placed randomly among the non-observed regions. In the near miss hidden EZ scenario, the EZs are placed among the non-observed nodes that are well-connected to at least three observed nodes, where well-connected means having the outgoing connection stronger that 97-percentile of all connections weights. Thirty-two synthetic seizures were generated and fitted for each scenario. (B) Precision-recall curves obtained by using the inferred probability *p*(*c > c*_*h*_) that a region is highly excitable as a predictor and varying a threshold *p*_*t*_ for this predictor from zero to one. Two specific thresholds *p*_*t*_ are marked on each curve.

---

**Figure S5:**
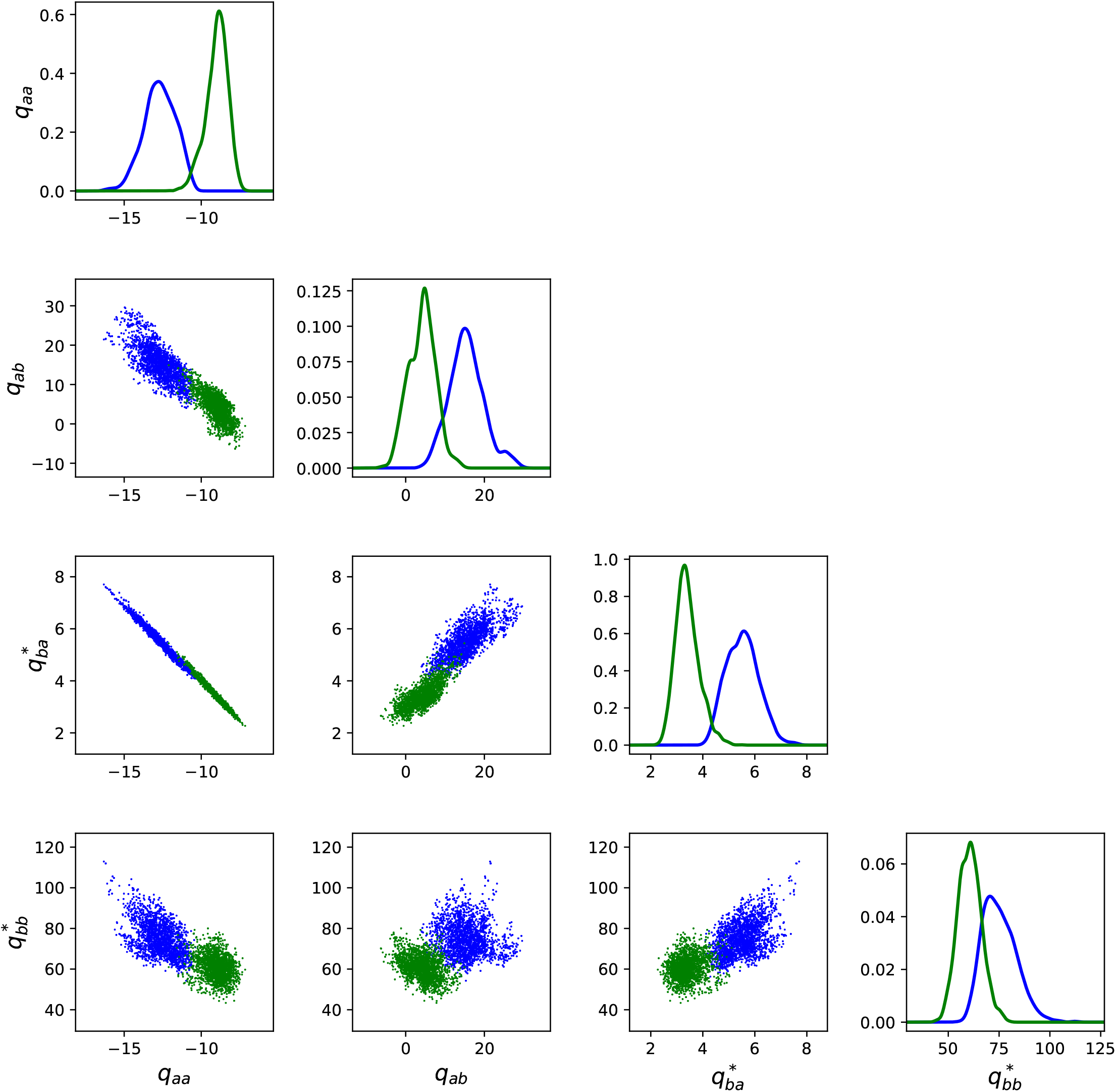
Posterior distribution of the hyperparameters. The diagonal panels show the kernel density estimation of the posterior distributions, the offdiagonal panels show the pair plots of the hyperparameters. The two colors represent two different data folds.

**Figure S6:**
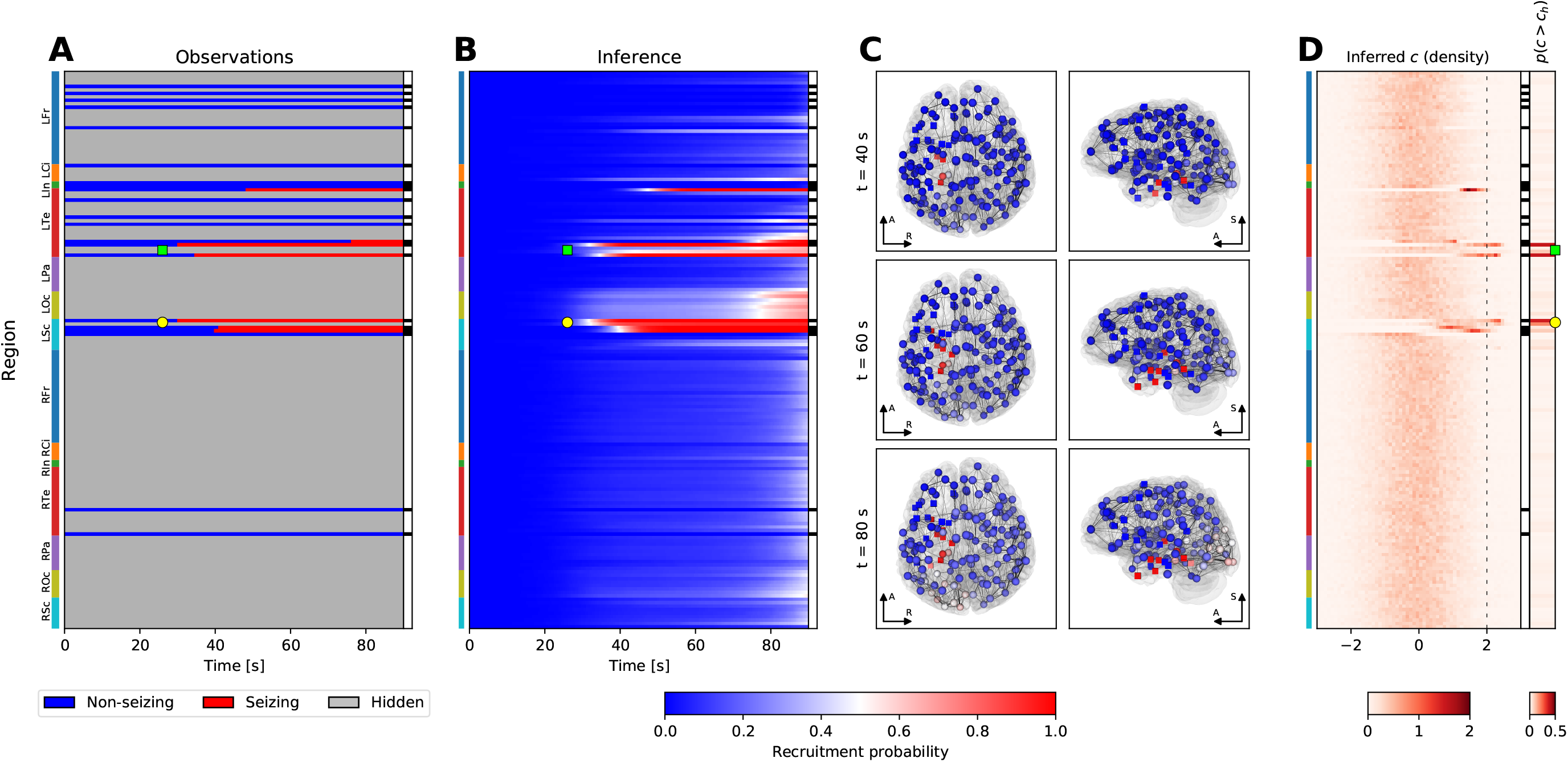
Example of the inference results on a seizure from subject 17. The layout of the figure is the same as in Fig. 5. The seizure is observed to start in the left anterior hippocampus (yellow circle, panels A, B) and left rhinal cortex and collateral sulcus (green square, panels A, B), and then it spreads to left amygdala and thalamus, left temporal pole, and eventually to left occipito-temporal sulcus. The seizure activity is inferred to remain spatially restricted, only the possible late recruitment of the left occipital lobe is inferred in addition to the observations. The early seizing regions are also inferred as being epileptogenic (yellow circle and green square, panel D).

**Figure S7:**
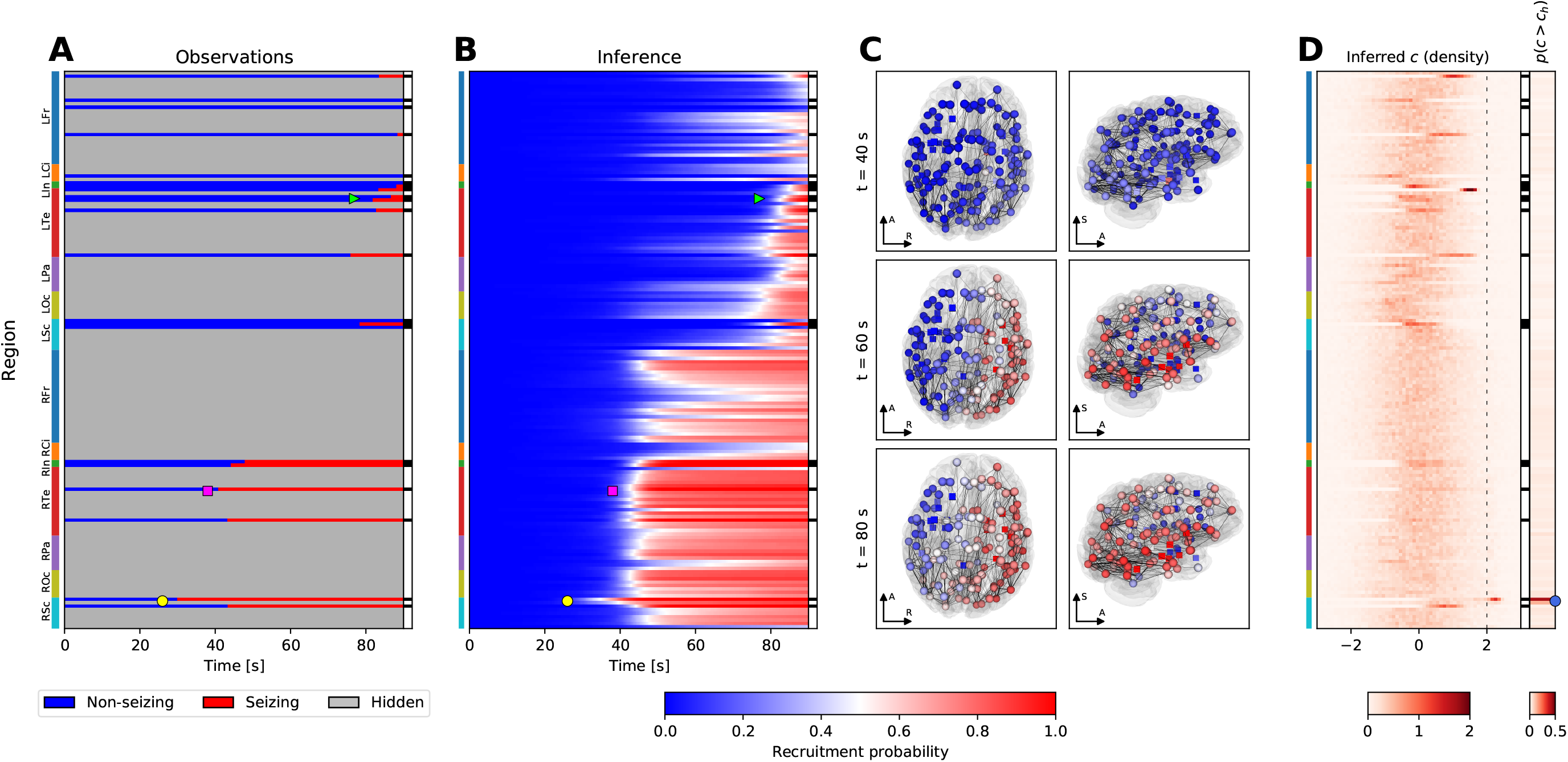
Example of the inference results on a seizure from subject 1. The layout of the figure is the same as in Fig. 5. From the observations the method infers that the seizure starts in the anterior part of the right hippocampus (yellow circle, panels A, B) before a large portion of the right hemisphere is recruited (magenta square, panels A, B), and eventually also some regions in the left hemisphere (light green triangle, panels A, B). The anterior part of the right hippocampus is strongly inferred to be the epileptogenic zone; the inference also points to the strongly connected posterior part of the right hippocampus (blue circle, panel D).

**Figure S8:**
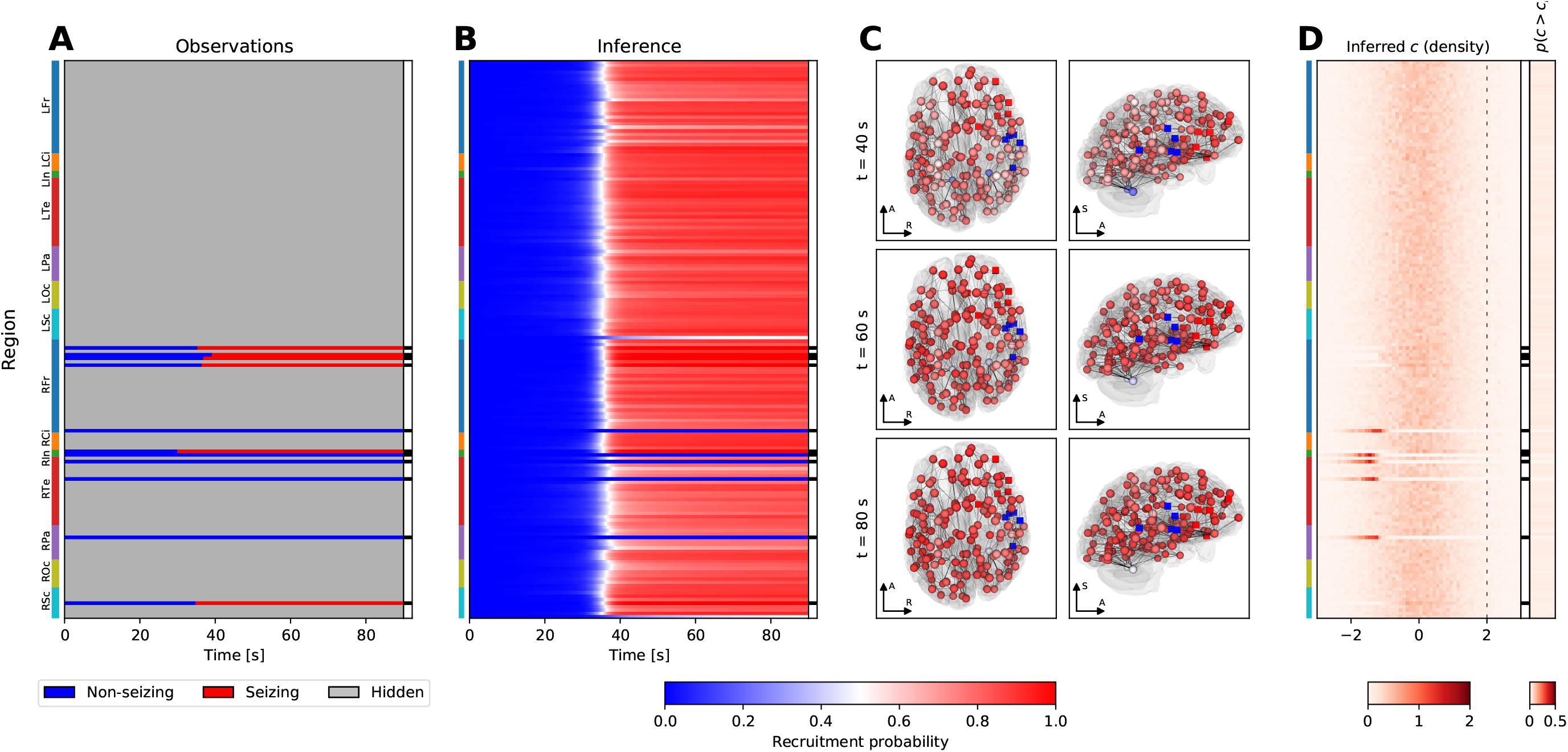
Example of the inference results on a seizure from subject 36. The layout of the figure is the same as in Fig. 5. This example shows a possible failure of the model, with a pattern repeatedly occurring among the results. Even though several stable regions are observed during the seizure, the method infers almost simultaneous recruitment of the majority of brain regions. The only regions not recruited are the observed stable nodes, and the left and right cerebella which are in the model generally very stable regions due to the low volumetric density of afferent projections obtained from diffusion-weighted imaging. None of the regions is identified as more epileptogenic than others; in cases of simulateneous recruitment it is difficult to infer which region is leading the seizure activity and which regions are only following.

**Figure S9:**
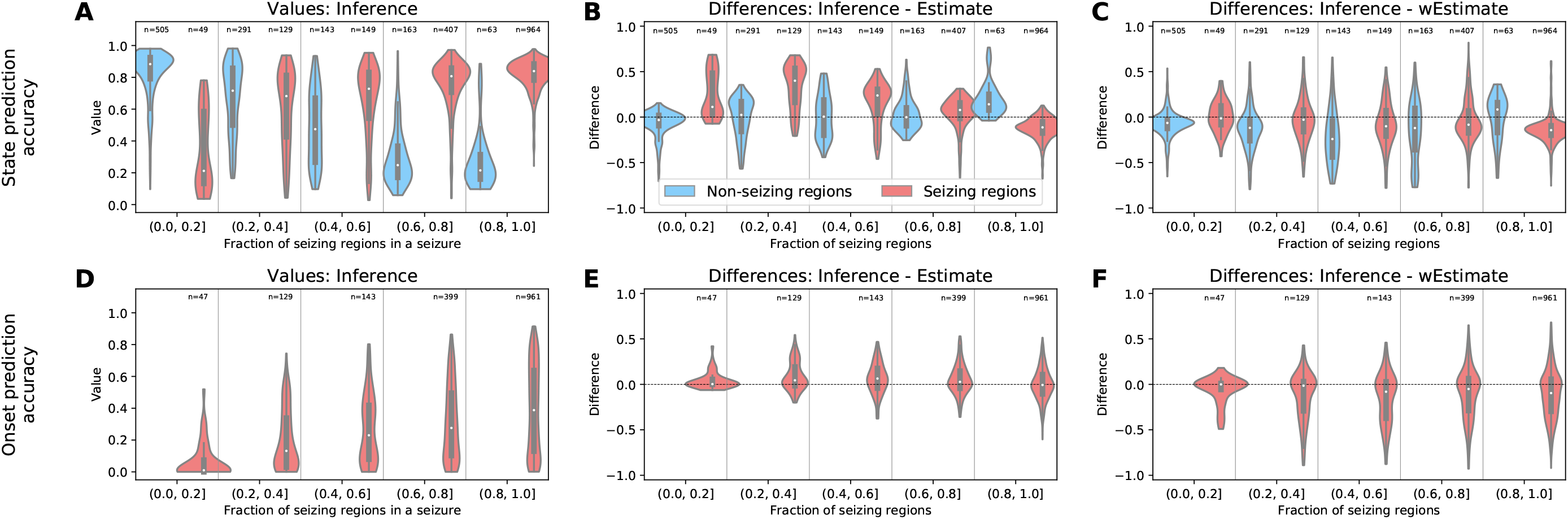
Results of the leave-one-out cross-validation for patient data. Top row: results for the state prediction accuracy, bottom row: results for the onset prediction accuracy. The onset prediction accuracy is evaluated only for the seizing regions. (A, D) Values of the accuracies obtained by the inference for the seizing and non-seizing regions, divided based on the fraction of seizing regions among the observed regions in a seizure. Each datapoint from which the violin plots are constructed corresponds to the accuracy value for a single region. (B, E) Differences between the accuracy values obtained by the inference and the unweighted estimate. The layout is the same as in panels A and D. (C, F) Differences between the accuracy values obtained by the inference and the weighted estimate. The layout is the same as in panels A and D. In all panels, the violin plots show a kernel density estimate of a given variable. The inner boxplots show the median (white dot), interquartile range (IQR, gray bar) and adjacent values (upper/lower quartile +/−1.5 IQR, gray line).

**Table S1:** List of operations to create the custom brain parcellation. The operations are: `merge` -merges two or more regions together; `rename` -renames a region; `split` -splits one region into multiple using a linear PCA projection of the coordinates; `split-to` -splits one region into multiple using a linear projection and merges the created regions with the specified existing ones; `split-to-nl` -splits one region into multiple using a nonlinear Isomap projection and merges the created regions with the specified existing ones; `split-mes-`splits the region into the mesial wall and the remaining part, using a criterion of the mesial-lateral component of the unit normal vector of the inflated cortical surface being equal to −0.5. Wildcards %h and %H stand for the hemisphere name in the format ‘lh’/’rh’ and ’Left’/’Right’ respectively, and numerical wildcards %1, %2, %3 stand for temporary regions which are then replaced by following operations. If ratios are given, the split is into non-equal sized region as determined by the ratios, otherwise the region is split into equally sized regions. For the cortical regions, the operations are performed on the triangulated cortical surface, while for the subcortical regions the operations are performed on the voxels.

| Operation | Name(s) of the input region(s) | Name(s) of the output region(s) | Options |
| --- | --- | --- | --- |
| merge | ctx_%h_G_and_S_frontomargin, ctx_%h_G_and_S_transv_frontopol | %H-Frontal-pole |  |
| merge | ctx_%h_G_orbital, ctx_%h_S_orbital_lateral, ctx_%h_S_orbital-H_Shaped | %H-Orbito-frontal-cortex |  |
| merge | ctx_%h_G_rectus, ctx_%h_S_suborbital, ctx_%h_S_orbital_med-olfact | %H-Gyrus-rectus |  |
| merge | ctx_%h_G_front_inf-Orbital, ctx_%h_Lat_Fis-ant-Horizont | %H-F3-Pars-Orbitalis |  |
| merge | ctx_%h_G_front_inf-Triangul, ctx_%h_Lat_Fis-ant-Vertical | %H-F3-Pars-triangularis |  |
| merge | ctx_%h_G_temp_sup-G_T_transv, ctx_%h_S_temporal_transverse | %H-Gyrus-of-Heschl |  |
| merge | ctx_%h_Lat_Fis-post, ctx_%h_G_temp_sup-Plan_tempo | %H-T1-planum-temporale |  |
| merge | ctx_%h_G_occipital_sup, ctx_%h_S_oc_sup_and_transversal | %H-O1 |  |
| merge | ctx_%h_G_precuneus, ctx_%h_S_subparietal | %H-Precuneus |  |
| merge | ctx_%h_G_occipital_middle, ctx_%h_S_oc_middle_and_Lunatus | %H-O2 |  |
| rename | ctx_%h_G_front_inf-Opercular | %H-F3-pars-opercularis |  |
| rename | ctx_%h_S_front_inf | %H-Inferior-frontal-sulcus |  |
| rename | ctx_%h_S_front_middle | %H-Middle-frontal-sulcus |  |
| rename | ctx_%h_G_subcallosal | %H-Subcallosal-area |  |
| rename | ctx_%h_S_precentral-inf-part | %H-Precentral-sulcus-inferior-part |  |
| rename | ctx_%h_S_precentral-sup-part | %H-Precentral-sulcus-superior-part |  |
| rename | ctx_%h_G_and_S_paracentral | %H-Paracentral-lobule |  |
| rename | ctx_%h_Pole_temporal | %H-Temporal-pole |  |
| rename | ctx_%h_G_temp_sup-Plan_polar | %H-T1-planum-polare |  |
| rename | ctx_%h_G_oc-temp_lat-fusifor | %H-Fusiform-gyrus |  |
| rename | ctx_%h_G_postcentral | %H-Postcentral-gyrus |  |
| rename | ctx_%h_S_postcentral | %H-Postcentral-sulcus |  |
| rename | ctx_%h_G_parietal_sup | %H-Superior-parietal-lobule-P1 |  |
| rename | ctx_%h_G_pariet_inf-Angular | %H-Angular-gyrus |  |
| rename | ctx_%h_S_intrapariet_and_P_trans | %H-Intraparietal-sulcus |  |
| rename | ctx_%h_S_cingul-Marginalis | %H-Marginal-branch-of-the-cingulate-sulcus |  |
| rename | ctx_%h_S_parieto_occipital | %H-Parieto-occipital-sulcus |  |
| rename | ctx_%h_S_occipital_ant | %H-Anterior-occipital-sulcus-and-preoccipital-notch |  |
| rename | ctx_%h_G_and_S_occipital_inf | %H-O3 |  |
| rename | ctx_%h_Pole_occipital | %H-Occipital-pole |  |
| rename | ctx_%h_G_oc-temp_med-Lingual | %H-Lingual-gyrus |  |
| rename | ctx_%h_S_calcarine | %H-Calcarine-sulcus |  |
| rename | ctx_%h_G_cuneus | %H-Cuneus |  |
| split | ctx_%h_G_front_middle | %H-F2-rostral, %H-F2-caudal |  |
| split | ctx_%h_S_front_sup | %H-SFS-rostral, %H-SFS-caudal |  |
| split | ctx_%h_G_and_S_subcentral | %H-Central-operculum, %H-Parietal-operculum |  |
| split | ctx_%h_G_temp_sup-Lateral | %H-T1-lateral-anterior, %H-T1-lateral-posterior |  |
| split | ctx_%h_S_temporal_sup | %H-STS-anterior, %H-STS-posterior |  |
| split | ctx_%h_S_temporal_inf | %H-ITS-anterior, %H-ITS-posterior |  |
| split | ctx_%h_G_temporal_middle | %H-T2-anterior, %H-T2-posterior |  |
| split | ctx_%h_G_temporal_inf | %H-T3-anterior, %H-T3-posterior |  |
| split-mes | ctx_%h_G_front_sup | %1, %2 |  |
| split | %1 | %H-F1-mesial-prefrontal, %H-PreSMA, %H-SMA | ratios: 2, 1, 3 |
| split | %2 | %H-F1-lateral-prefrontal, %H-F1-lateral-premotor |  |
| split | ctx_%h_G_precentral | %H-Precentral-gyrus-head-face, %H-Precentral-gyrus-upper-limb | ratios: 2, 1 |
| split | ctx_%h_S_central | %H-Central-sulcus-head-face, %H-Central-sulcus-upper-limb | ratios: 2, 1 |
| split | ctx_%h_S_oc-temp_med_and_Lingual | %H-Collateral-sulcus, %1 |  |
| merge | %1, ctx_%h_S_collat_transv_post | %H-Lingual-sulcus |  |
| split | ctx_%h_G_oc-temp_med-Parahip | %1, %H-Parahippocampal-cortex |  |
| split | ctx_%h_S_collat_transv_ant | %2, %3 |  |
| merge | %1, %2 | %H-Rhinal-cortex |  |
| merge | %3, ctx_%h_S_oc-temp_lat | %H-Occipito-temporal-sulcus |  |
| split | ctx_%h_G_pariet_inf-Supramar | %H-Supramarginal-anterior, %1 |  |
| merge | %1, ctx_%h_S_interm_prim-Jensen | %H-Supramarginal-posterior |  |
| split-to-nl | ctx_%h_S_pericallosal | ctx_%h_G_and_S_cingul-Ant, ctx_%h_G_and_S_cingul-Mid-Ant, ctx_%h_G_and_S_cingul-Mid-Post, ctx_%h_G_cingul-Post-dorsal, ctx_%h_G_cingul-Post-ventral |  |
| rename | ctx_%h_G_and_S_cingul-Ant | %H-Anterior-cingulate-cortex |  |
| rename | ctx_%h_G_and_S_cingul-Mid-Ant | %H-Middle-cingulate-cortex-anterior-part |  |
| rename | ctx_%h_G_and_S_cingul-Mid-Post | %H-Middle-cingulate-cortex-posterior-part |  |
| rename | ctx_%h_G_cingul-Post-dorsal | %H-Posterior-cingulate-cortex-dorsal |  |
| rename | ctx_%h_G_cingul-Post-ventral | %H-Posterior-cingulate-cortex-retrosplenial-gyrus |  |
| split-to | ctx_%h_S_circular_insula_sup | ctx_%h_G_insular_short, ctx_%h_G_Ins_lg_and_S_cent_ins |  |
| merge | ctx_%h_S_circular_insula_ant, ctx_%h_G_insular_short | %H-Insula-gyri-brevi |  |
| merge | ctx_%h_G_Ins_lg_and_S_cent_ins, ctx_%h_S_circular_insula_inf | %H-Insula-gyri-longi |  |
| split | %H-Hippocampus | %H-Hippocampus-anterior, %H-Hippocampus-posterior |  |
| rename | %H-Cerebellum-Cortex | %H-Cerebellar-cortex |  |
| rename | %H-Thalamus-Proper | %H-Thalamus |  |
| rename | %H-Caudate | %H-Caudate-nucleus |  |
| rename | %H-Accumbens-area | %H-Nucleus-accumbens |  |

**Table S2:** List of brain regions in the custom atlas.

| Anatomical grouping | Index | Name | Index | Name |
| --- | --- | --- | --- | --- |
| Frontal lobe | 1 | Left-Frontal-pole | 82 | Right-Frontal-pole |
| Frontal lobe | 2 | Left-Orbito-frontal-cortex | 83 | Right-Orbito-frontal-cortex |
| Frontal lobe | 3 | Left-Gyrus-rectus | 84 | Right-Gyrus-rectus |
| Frontal lobe | 4 | Left-F3-Pars-Orbitalis | 85 | Right-F3-Pars-Orbitalis |
| Frontal lobe | 5 | Left-F3-Pars-triangularis | 86 | Right-F3-Pars-triangularis |
| Frontal lobe | 6 | Left-F3-pars-opercularis | 87 | Right-F3-pars-opercularis |
| Frontal lobe | 7 | Left-Inferior-frontal-sulcus | 88 | Right-Inferior-frontal-sulcus |
| Frontal lobe | 8 | Left-F2-rostral | 89 | Right-F2-rostral |
| Frontal lobe | 9 | Left-F2-caudal | 90 | Right-F2-caudal |
| Frontal lobe | 10 | Left-Middle-frontal-sulcus | 91 | Right-Middle-frontal-sulcus |
| Frontal lobe | 11 | Left-SFS-rostral | 92 | Right-SFS-rostral |
| Frontal lobe | 12 | Left-SFS-caudal | 93 | Right-SFS-caudal |
| Frontal lobe | 13 | Left-F1-mesial-prefrontal | 94 | Right-F1-mesial-prefrontal |
| Frontal lobe | 14 | Left-PreSMA | 95 | Right-PreSMA |
| Frontal lobe | 15 | Left-SMA | 96 | Right-SMA |
| Frontal lobe | 16 | Left-F1-lateral-prefrontal | 97 | Right-F1-lateral-prefrontal |
| Frontal lobe | 17 | Left-F1-lateral-premotor | 98 | Right-F1-lateral-premotor |
| Frontal lobe | 18 | Left-Subcallosal-area | 99 | Right-Subcallosal-area |
| Frontal lobe | 19 | Left-Precentral-sulcus-inferior-part | 100 | Right-Precentral-sulcus-inferior-part |
| Frontal lobe | 20 | Left-Precentral-sulcus-superior-part | 101 | Right-Precentral-sulcus-superior-part |
| Frontal lobe | 21 | Left-Precentral-gyrus-head-face | 102 | Right-Precentral-gyrus-head-face |
| Frontal lobe | 22 | Left-Precentral-gyrus-upper-limb | 103 | Right-Precentral-gyrus-upper-limb |
| Frontal lobe | 23 | Left-Central-sulcus-head-face | 104 | Right-Central-sulcus-head-face |
| Frontal lobe | 24 | Left-Central-sulcus-upper-limb | 105 | Right-Central-sulcus-upper-limb |
| Frontal lobe | 25 | Left-Paracentral-lobule | 106 | Right-Paracentral-lobule |
| Frontal lobe | 26 | Left-Central-operculum | 107 | Right-Central-operculum |
| Frontal lobe | 27 | Left-Parietal-operculum | 108 | Right-Parietal-operculum |
| Cingulate cortex | 28 | Left-Anterior-cingulate-cortex | 109 | Right-Anterior-cingulate-cortex |
| Cingulate cortex | 29 | Left-Middle-cingulate-cortex-anterior-part | 110 | Right-Middle-cingulate-cortex-anterior-part |
| Cingulate cortex | 30 | Left-Middle-cingulate-cortex-posterior-part | 111 | Right-Middle-cingulate-cortex-posterior-part |
| Cingulate cortex | 31 | Left-Posterior-cingulate-cortex-dorsal | 112 | Right-Posterior-cingulate-cortex-dorsal |
| Cingulate cortex | 32 | Left-Posterior-cingulate-cortex-retrosplenial-gyrus | 113 | Right-Posterior-cingulate-cortex-retrosplenial-gyrus |
| Insula | 33 | Left-Insula-gyri-brevi | 114 | Right-Insula-gyri-brevi |
| Insula | 34 | Left-Insula-gyri-longi | 115 | Right-Insula-gyri-longi |
| Temporal lobe | 35 | Left-Temporal-pole | 116 | Right-Temporal-pole |
| Temporal lobe | 36 | Left-T1-planum-polare | 117 | Right-T1-planum-polare |
| Temporal lobe | 37 | Left-Gyrus-of-Heschl | 118 | Right-Gyrus-of-Heschl |
| Temporal lobe | 38 | Left-T1-planum-temporale | 119 | Right-T1-planum-temporale |
| Temporal lobe | 39 | Left-T1-lateral-anterior | 120 | Right-T1-lateral-anterior |
| Temporal lobe | 40 | Left-T1-lateral-posterior | 121 | Right-T1-lateral-posterior |
| Temporal lobe | 41 | Left-STS-anterior | 122 | Right-STS-anterior |
| Temporal lobe | 42 | Left-STS-posterior | 123 | Right-STS-posterior |
| Temporal lobe | 43 | Left-ITS-anterior | 124 | Right-ITS-anterior |
| Temporal lobe | 44 | Left-ITS-posterior | 125 | Right-ITS-posterior |
| Temporal lobe | 45 | Left-T2-anterior | 126 | Right-T2-anterior |
| Temporal lobe | 46 | Left-T2-posterior | 127 | Right-T2-posterior |
| Temporal lobe | 47 | Left-T3-anterior | 128 | Right-T3-anterior |
| Temporal lobe | 48 | Left-T3-posterior | 129 | Right-T3-posterior |
| Temporal lobe | 49 | Left-Fusiform-gyrus | 130 | Right-Fusiform-gyrus |
| Temporal lobe | 50 | Left-Occipito-temporal-sulcus | 131 | Right-Occipito-temporal-sulcus |
| Temporal lobe | 51 | Left-Collateral-sulcus | 132 | Right-Collateral-sulcus |
| Temporal lobe | 52 | Left-Lingual-sulcus | 133 | Right-Lingual-sulcus |
| Temporal lobe | 53 | Left-Parahippocampal-cortex | 134 | Right-Parahippocampal-cortex |
| Temporal lobe | 54 | Left-Rhinal-cortex | 135 | Right-Rhinal-cortex |
| Parietal lobe | 55 | Left-Postcentral-gyrus | 136 | Right-Postcentral-gyrus |
| Parietal lobe | 56 | Left-Postcentral-sulcus | 137 | Right-Postcentral-sulcus |
| Parietal lobe | 57 | Left-Superior-parietal-lobule-P1 | 138 | Right-Superior-parietal-lobule-P1 |
| Parietal lobe | 58 | Left-Supramarginal-anterior | 139 | Right-Supramarginal-anterior |
| Parietal lobe | 59 | Left-Supramarginal-posterior | 140 | Right-Supramarginal-posterior |
| Parietal lobe | 60 | Left-Angular-gyrus | 141 | Right-Angular-gyrus |
| Parietal lobe | 61 | Left-Intraparietal-sulcus | 142 | Right-Intraparietal-sulcus |
| Parietal lobe | 62 | Left-Precuneus | 143 | Right-Precuneus |
| Parietal lobe | 63 | Left-Marginal-branch-of-the-cingulate-sulcus | 144 | Right-Marginal-branch-of-the-cingulate-sulcus |
| Parietal lobe | 64 | Left-Parieto-occipital-sulcus | 145 | Right-Parieto-occipital-sulcus |
| Occipital lobe | 65 | Left-Anterior-occipital-sulcus-and-preoccipital-notch | 146 | Right-Anterior-occipital-sulcus-and-preoccipital-notch |
| Occipital lobe | 66 | Left-O3 | 147 | Right-O3 |
| Occipital lobe | 67 | Left-O2 | 148 | Right-O2 |
| Occipital lobe | 68 | Left-O1 | 149 | Right-O1 |
| Occipital lobe | 69 | Left-Occipital-pole | 150 | Right-Occipital-pole |
| Occipital lobe | 70 | Left-Lingual-gyrus | 151 | Right-Lingual-gyrus |
| Occipital lobe | 71 | Left-Calcarine-sulcus | 152 | Right-Calcarine-sulcus |
| Occipital lobe | 72 | Left-Cuneus | 153 | Right-Cuneus |
| Subcortical regions | 73 | Left-Hippocampus-anterior | 154 | Right-Hippocampus-anterior |
| Subcortical regions | 74 | Left-Hippocampus-posterior | 155 | Right-Hippocampus-posterior |
| Subcortical regions | 75 | Left-Amygdala | 156 | Right-Amygdala |
| Subcortical regions | 76 | Left-Thalamus | 157 | Right-Thalamus |
| Subcortical regions | 77 | Left-Caudate-nucleus | 158 | Right-Caudate-nucleus |
| Subcortical regions | 78 | Left-Putamen | 159 | Right-Putamen |
| Subcortical regions | 79 | Left-Pallidum | 160 | Right-Pallidum |
| Subcortical regions | 80 | Left-Nucleus-accumbens | 161 | Right-Nucleus-accumbens |
| Subcortical regions | 81 | Left-Cerebellar-cortex | 162 | Right-Cerebellar-cortex |

**Table S3:** Patient table. Abbreviations: AVM, arteriovenous malformation; DNET, dysembryoplastic neuroepithelial tumor; FCD, focal cortical dyplasia; HH, hypothalamic hamartoma; L, left; NA, not applicable; PMG, polymicrogyria; PNH, periventricular nodular heterotopia; R, right. The column “Number of seizures” shows the number of recorded seizures longer than 30 seconds used in this study.

| Patient | Gender | Age at epilepsy onset (y) | Epilepsy duration (y) | Epilepsy type | MRI | Histopathology | Side | Engel score | Post-op MRI | Number of seizures |
| --- | --- | --- | --- | --- | --- | --- | --- | --- | --- | --- |
| 1 | F | 31 | 3 | Temporo-insular | Normal | Hippocampal sclerosis | R | I | Y | 4 |
| 2 | F | 19 | 10 | Temporo-occipital | L temporo-occipital PNH | NA | L | NA | N | 3 |
| 3 | M | 23 | 13 | Temporo-frontal | R temporo-occipital scar | FCD1a | R | I | Y | 3 |
| 4 | F | 23 | 3 | Temporal | R temporal mesial ganglioglioma | Ganglioglioma | R | I | Y | 2 |
| 5 | M | 0.3 | 21 | Postcentral - superior parietal | L postcentral-parietal gyration asymmetry | NA | L | NA | N | 1 |
| 6 | M | 45 | 14 | Fronto-temporal | Normal | NA | L | NA | N | 3 |
| 7 | M | 55 | 5 | Temporal | Normal | Slight gliosis | R>L | III | Y | 2 |
| 8 | F | 38 | 8 | Temporal | L amygdala enlargement | Slight gliosis | L | III | N | 3 |
| 9 | F | 11 | 34 | Bifocal: parietal mesial & temporo-basal | Unknown R parietal lesion | Rosenthal fibers; slight gliosis | R | III | Y | 2 |
| 10 | F | 27 | 18 | Temporal | L hippocampal sclerosis | Hippocampal sclerosis | L | I | N | 6 |
| 11 | F | 27 | 14 | Frontal | L frontal scar (abcess) | Gliosis | L | IV | Y | 3 |
| 12 | F | 19 | 9 | Bilateral temporo-frontal | Bilateral hippocampal & amygdala T2-hypersignal | NA | R&L | NA | N | 2 |
| 13 | M | 2 | 17 | Frontal | Normal | Slight gliosis | L | I | N | 2 |
| 14 | F | 5 | 18 | Premotor | Normal | FCD2b | L | I | Y | 0 |
| 15 | M | 8 | 33 | Temporal | R temporal PMG & multiple PNH | NA | R | NA | N | 2 |
| 16 | M | 6 | 23 | Temporo-operculo-fronto-parietal | R temporo-parieto-insular & L temporo-parietal necrosis | NA | R>L | NA | N | 4 |
| 17 | M | 5 | 21 | Temporal | L temporo-polar hypothyrophy and hippocampal sclerosis | Hippocampal sclerosis; gliosis | L | I | Y | 4 |
| 18 | M | 2 | 22 | Parieto-temporal | L Parieto-occipital necrosis (perinatal anoxia) | NA | L | NA | N | 2 |
| 19 | M | 29 | 15 | Temporo-insular | Normal | NA | L>R | NA | N | 5 |
| 20 | F | 17 | 10 | Temporal | Normal | Hippocampal sclerosis | R | I | Y | 3 |
| 21 | F | 9 | 14 | Occipital | Normal | FCD1c | L | II | Y | 2 |
| 22 | F | 7 | 23 | Parietal | L parietal FCD | FCD2b | L | I | Y | 0 |
| 23 | M | 35 | 28 | Temporal | Normal | NA | L | NA | N | 2 |
| 24 | M | 14 | 15 | Temporal | Normal | NA | R | NA | N | 4 |
| 25 | M | 7 | 35 | Insular | Normal | NA | L | I | Y | 4 |
| 26 | F | 4 | 24 | Occipital | PNH | NA | R | NA | N | 0 |
| 27 | M | 17 | 12 | Frontal | R prefrontal gliotic scar (AVM) | Gliosis | R>L | II | N | 3 |
| 28 | F | 8 | 14 | Temporo-frontal | Anterior temporal necrosis | Gliosis | R | III | Y | 3 |
| 29 | F | 21 | 9 | Bilateral temporal | Bilateral posterior PNH | NA | R>L | NA | N | 3 |

| Patient | Gender | Age at<br>epilepsy<br>onset (y) | Epilepsy<br>duration<br>(y) | Epilepsy type | MRI | Histopathology | Side | Engel<br>score | Post-op<br>MRI | Number<br>of<br>seizures |
| --- | --- | --- | --- | --- | --- | --- | --- | --- | --- | --- |
| 30 | M | 11 | 45 | Temporo-frontal | R Frontal FCD | FCD 2 | R | I | Y | 1 |
| 31 | F | 20 | 18 | Occipital | Normal | NA | R | NA | N | 3 |
| 32 | F | 15 | 21 | Bilateral temporal with HH | L HH | NA | R&L | NA | N | 5 |
| 33 | F | 18 | 5 | Temporo-parieto-opercular | Normal | Hippocampal sclerosis | R | IV | Y | 9 |
| 34 | F | 33 | 8 | Temporal | Multiple R temporo-parietal<br>PNH & temporal PMG | NA | R | NA | N | 2 |
| 35 | M | 4 | 27 | Bilateral occipito-temporal | R occipital mesial FCD | NA | R&L | NA | N | 4 |
| 36 | F | 8 | 13 | Temporo-insular | R temporal anterior resection<br>cavity | Gliosis | R | IV | Y | 3 |
| 37 | M | 28 | 5 | Temporal mesial | R temporo-polar & amygdala<br>FCD, L post-chiasmal pilocytic<br>astrocytoma | FCD 2b | R | III | Y | 4 |
| 38 | M | 11 | 22 | Bilateral temporal | Normal | NA | L&R | NA | N | 8 |
| 39 | M | 40 | 4 | Temporo-frontal | R fronto-temporal necrosis<br>(gunshot injury) | Gliosis | R | I | Y | 3 |
| 40 | F | 16 | 19 | Temporal mesial | Hippocampal sclerosis | Hippocampal sclerosis | L | II | Y | 2 |
| 41 | M | 0.7 | 26 | Bilateral, temporal predominant | R perisylvian necrosis (perinatal<br>stroke) | NA | R>L | NA | N | 4 |
| 42 | F | 9 | 19 | Temporal mesial | Bilateral hippocampal sclerosis | NA | L | NA | N | 6 |
| 43 | F | 7 | 16 | Premotor | R precentral FCD | NA | R | NA | N | 1 |
| 44 | M | 0.5 | 39 | Multifocal:<br>parieto-operculo-premotor;<br>temporal mesial | L hippocampal & amygdala T2<br>hypersignal | NA | L | NA | N | 2 |
| 45 | F | 24 | 17 | Temporal mesial | Normal | NA | L | IV | Y | 3 |
| 46 | M | 1.5 | 31 | Insulo-parieto-premotor | Normal | NA | R | NA | N | 0 |
| 47 | M | 16 | 13 | Bilateral frontal | Normal | NA | R&L | NA | N | 2 |
| 48 | F | 15 | 7 | Premotor | R parietal DNET | NA | R | NA | N | 1 |
| 49 | F | 1 | 21 | Motor-opercular | R fronto-opercular resection<br>cavity | NA | R | NA | N | 1 |
| 50 | M | 14 | 21 | Motor-premotor | L insulo-opercular necrosis<br>(stroke) | NA | L | I | Y | 0 |

## References

Aguirre, L. A., Portes, L. L., Letellier, C., Oct. 2018. Structural, dynamical and symbolic observability: From dynamical systems to networks. PLOS ONE 13 (10), e0206180.

An, S., Bartolomei, F., Guye, M., Jirsa, V., Jun. 2019. Optimization of surgical intervention outside the epileptogenic zone in the Virtual Epileptic Patient (VEP). PLOS Computational Biology 15 (6), e1007051.

Aubert, S., Wendling, F., Regis, J., McGonigal, A., Figarella-Branger, D., Peragut, J.-C., Girard, N., Chauvel, P., Bartolomei, F., Nov. 2009. Local and remote epileptogenicity in focal cortical dysplasias and neurodevelopmental tumours. Brain 132 (11), 3072–3086.

Bartolomei, F., Chauvel, P., Wendling, F., Jul. 2008. Epileptogenicity of brain structures in human temporal lobe epilepsy: A quantified study from intracerebral EEG. Brain 131 (7), 1818–1830.

Bartolomei, F., Lagarde, S., Wendling, F., McGonigal, A., Jirsa, V., Guye, M., Bénar, C., May 2017. Defining epileptogenic networks: Contribution of SEEG and signal analysis. Epilepsia 58 (7), 1131–1147.

Baud, M. O., Perneger, T., Rácz, A., Pensel, M. C., Elger, C., Rydenhag, B., Malmgren, K., Cross, J. H., McKenna, G., Tisdall, M., Lamberink, H. J., Rheims, S., Ryvlin, P., Isnard, J., Mauguière, F., Arzimanoglou, A., Akkol, S., Deniz, K., Ozkara, C., Lossius, M., Rektor, I., Kälviäinen, R., Vanhatalo, L.-M., Dimova, P., Minkin, K., Staack, A. M., Steinhoff, B. J., Kalina, A., Krsek, P., Marusic, P., Jordan, Z., Fabo, D., Carrette, E., Boon, P., Rocka, S., Mameniškien.e, R., Vulliemoz, S., Pittau, F., Braun, K. P. J., Seeck, M., Jun. 2018. European trends in epilepsy surgery. Neurology 91 (2), e96–e106.

Besson, P., Bandt, S. K., Proix, T., Lagarde, S., Jirsa, V. K., Ranjeva, J.-P., Bartolomei, F., Guye, M., Oct. 2017. Anatomic consistencies across epilepsies: A stereotactic-EEG informed high-resolution structural connectivity study. Brain 140 (10), 2639–2652.

Betancourt, M., Jan. 2017. A Conceptual Introduction to Hamiltonian Monte Carlo.

Burns, S. P., Santaniello, S., Yaffe, R. B., Jouny, C. C., Crone, N. E., Bergey, G. K., Anderson, W. S., Sarma, S. V., Nov. 2014. Network dynamics of the brain and influence of the epileptic seizure onset zone. Proceedings of the National Academy of Sciences 111 (49), E5321–E5330.

Cam, S. L., Ranta, R., Caune, V., Korats, G., Koessler, L., Maillard, L., Louis-Dorr, V., Jun. 2017. SEEG dipole source localization based on an empirical Bayesian approach taking into account forward model uncertainties. NeuroImage 153, 1–15.

Carpenter, B., Gelman, A., Hoffman, M. D., Lee, D., Goodrich, B., Betancourt, M., Brubaker, M., Guo, J., Li, P., Riddell, A., 2017. Stan: A Probabilistic Programming Language. Journal of Statistical Software 76 (1).

Caune, V., Ranta, R., Le Cam, S., Hofmanis, J., Maillard, L., Koessler, L., Louis-Dorr, V., Sep. 2014. Evaluating dipolar source localization feasibility from intracerebral SEEG recordings. NeuroImage 98, 118–133.

Cooray, G. K., Sengupta, B., Douglas, P. K., Friston, K., Jan. 2016. Dynamic causal modelling of electrographic seizure activity using Bayesian belief updating. NeuroImage 125, 1142–1154.

Cover, T. M., Thomas, J. A., 2006. Elements of Information Theory, 2nd Edition. Wiley-Interscience, Hoboken, N.J.

Destrieux, C., Fischl, B., Dale, A., Halgren, E., Oct. 2010. Automatic parcellation of human cortical gyri and sulci using standard anatomical nomenclature. NeuroImage 53 (1), 1–15.

Devinsky, O., Vezzani, A., O’Brien, T. J., Jette, N., Scheffer, I. E., de Curtis, M., Perucca, P., May 2018. Epilepsy. Nature Reviews Disease Primers 4 (1), 1–24.

Feller, W., 1968. An Introduction to Probability Theory and Its Applications, 3rd Edition. Wiley Series in Probability and Mathematical Statistics. Wiley, S.l.

Fischl, B., Aug. 2012. FreeSurfer. NeuroImage 62 (2), 774–781.

Fischl, B., Salat, D. H., Busa, E., Albert, M., Dieterich, M., Haselgrove, C., van der Kouwe, A., Killiany, R., Kennedy, D., Klaveness, S., Montillo, A., Makris, N., Rosen, B., Dale, A. M., Jan. 2002. Whole Brain Segmentation: Automated Labeling of Neuroanatomical Structures in the Human Brain. Neuron 33 (3), 341–355.

Fisher, R. S., Scharfman, H. E., deCurtis, M., 2014. How Can We Identify Ictal and Interictal Abnormal Activity? In: Scharfman, H. E., Buckmaster, P. S. (Eds.), Issues in Clinical Epileptology: A View from the Bench. Advances in Experimental Medicine and Biology. Springer Netherlands, Dordrecht, pp. 3–23.

Freestone, D. R., Karoly, P. J., Nešić, D., Aram, P., Cook, M. J., Grayden, D. B., 2014. Estimation of effective connectivity via data-driven neural modeling. Frontiers in Neuroscience 8.

Gelman, A., Carlin, J. B., Stern, H. S., Dunson, D. B., Vehtari, A., Nov. 2013. Bayesian Data Analysis. Taylor & Francis Ltd.

Ghosh, A., Rho, Y., McIntosh, A. R., Kötter, R., Jirsa, V. K., Oct. 2008. Noise during Rest Enables the Exploration of the Brain’s Dynamic Repertoire. PLOS Computational Biology 4 (10), e1000196.

Goodfellow, M., Rummel, C., Abela, E., Richardson, M. P., Schindler, K., Terry, J. R., Jul. 2016. Estimation of brain network ictogenicity predicts outcome from epilepsy surgery. Scientific Reports 6 (1), 1–13.

Hashemi, M., Vattikonda, A. N., Sip, V., Guye, M., Bartolomei, F., Woodman, M. M., Jirsa, V. K., May 2020. The Bayesian Virtual Epileptic Patient: A probabilistic framework designed to infer the spatial map of epileptogenicity in a personalized large-scale brain model of epilepsy spread. NeuroImage, 116839.

Hebbink, J., Meijer, H., Huiskamp, G., van Gils, S., Leijten, F., 2017. Phenomenological network models: Lessons for epilepsy surgery. Epilepsia 58 (10), e147–e151.

Hoffman, M. D., Gelman, A., 2014. The No-U-Turn Sampler: Adaptively Setting Path Lengths in Hamiltonian Monte Carlo. Journal of Machine Learning Research 15, 1351–1381.

Honey, C. J., Sporns, O., Cammoun, L., Gigandet, X., Thiran, J. P., Meuli, R., Hagmann, P., Feb. 2009. Predicting human resting-state functional connectivity from structural connectivity. Proceedings of the National Academy of Sciences 106 (6), 2035–2040.

Hutchings, F., Han, C. E., Keller, S. S., Weber, B., Taylor, P. N., Kaiser, M., Dec. 2015. Predicting Surgery Targets in Temporal Lobe Epilepsy through Structural Connectome Based Simulations. PLOS Computational Biology 11 (12), e1004642.

Jayakar, P., Gotman, J., Harvey, A. S., Palmini, A., Tassi, L., Schomer, D., Dubeau, F., Bartolomei, F., Yu, A., Kršek, P., Velis, D., Kahane, P., Sep. 2016. Diagnostic utility of invasive EEG for epilepsy surgery: Indications, modalities, and techniques. Epilepsia 57 (11), 1735–1747.

Jehi, L., Yardi, R., Chagin, K., Tassi, L., Russo, G. L., Worrell, G., Hu, W., Cendes, F., Morita, M., Bartolomei, F., Chauvel, P., Najm, I., Gonzalez-Martinez, J., Bingaman, W., Kattan, M. W., Mar. 2015. Development and validation of nomograms to provide individualised predictions of seizure outcomes after epilepsy surgery: A retrospective analysis. The Lancet Neurology 14 (3), 283–290.

Jenkinson, M., Bannister, P., Brady, M., Smith, S., Oct. 2002. Improved Optimization for the Robust and Accurate Linear Registration and Motion Correction of Brain Images. NeuroImage 17 (2), 825–841.

Jenssen, S., Gracely, E. J., Sperling, M. R., 2006. How Long Do Most Seizures Last? A Systematic Comparison of Seizures Recorded in the Epilepsy Monitoring Unit. Epilepsia 47 (9), 1499–1503.

Jiménez-Jiménez, D., Nekkare, R., Flores, L., Chatzidimou, K., Bodi, I., Honavar, M., Mullatti, N., Elwes, R. D. C., Selway, R. P., Valentín, A., Alarcón, G., Feb. 2015. Prognostic value of intracranial seizure onset patterns for surgical outcome of the treatment of epilepsy. Clinical Neurophysiology 126 (2), 257–267.

Jirsa, V. K., Proix, T., Perdikis, D., Woodman, M. M., Wang, H., Gonzalez-Martinez, J., Bernard, C., Bénar, C., Guye, M., Chauvel, P., Bartolomei, F., Jan. 2017. The Virtual Epileptic Patient: Individualized whole-brain models of epilepsy spread. NeuroImage 145, 377–388.

Jirsa, V. K., Stacey, W. C., Quilichini, P. P., Ivanov, A. I., Bernard, C., Jul. 2014. On the nature of seizure dynamics. Brain 137 (8), 2110–2113.

Karoly, P. J., Kuhlmann, L., Soudry, D., Grayden, D. B., Cook, M. J., Freestone, D. R., Oct. 2018. Seizure pathways: A model-based investigation. PLOS Computational Biology 14 (10), e1006403.

Köster, J., Rahmann, S., Oct. 2012. Snakemake—a scalable bioinformatics workflow engine. Bioinformatics 28 (19), 2520–2522.

Kramer, M. A., Eden, U. T., Kolaczyk, E. D., Zepeda, R., Eskandar, E. N., Cash, S. S., Jul. 2010. Coalescence and Fragmentation of Cortical Networks during Focal Seizures. Journal of Neuroscience 30 (30), 10076–10085.

Lagarde, S., Bonini, F., McGonigal, A., Chauvel, P., Gavaret, M., Scavarda, D., Carron, R., Régis, J., Aubert, S., Villeneuve, N., Giusiano, B., Figarella-Branger, D., Trebuchon, A., Bartolomei, F., Jul. 2016. Seizure-onset patterns in focal cortical dysplasia and neurodevelopmental tumors: Relationship with surgical prognosis and neuropathologic subtypes. Epilepsia 57 (9), 1426–1435.

Laiou, P., Avramidis, E., Lopes, M. A., Abela, E., Müller, M., Akman, O. E., Richardson, M. P., Rummel, C., Schindler, K., Goodfellow, M., 2019. Quantification and Selection of Ictogenic Zones in Epilepsy Surgery. Frontiers in Neurology 10.

Liou, J.-y., Ma, H., Wenzel, M., Zhao, M., Baird-Daniel, E., Smith, E. H., Daniel, A., Emerson, R., Yuste, R., Schwartz, T. H., Schevon, C. A., Jul. 2018. Role of inhibitory control in modulating focal seizure spread. Brain 141 (7), 2083–2097.

Liou, J.-y., Smith, E. H., Bateman, L. M., Bruce, S. L., McKhann, G. M., Goodman, R. R., Emerson, R. G., Schevon, C. A., Abbott, L., Mar. 2020. A model for focal seizure onset, propagation, evolution, and progression. eLife 9, e50927.

Liu, Y.-Y., Barabási, A.-L., Sep. 2016. Control principles of complex systems. Reviews of Modern Physics 88 (3), 035006.

Lopes, M. A., Junges, L., Woldman, W., Goodfellow, M., Terry, J. R., 2020. The Role of Excitability and Network Structure in the Emergence of Focal and Generalized Seizures. Frontiers in Neurology 11.

Lopes, M. A., Richardson, M. P., Abela, E., Rummel, C., Schindler, K., Goodfellow, M., Terry,J. R., Aug. 2017. An optimal strategy for epilepsy surgery: Disruption of the rich-club? PLOS Computational Biology 13 (8), e1005637.

Lopes, M. A., Richardson, M. P., Abela, E., Rummel, C., Schindler, K., Goodfellow, M., Terry, J. R., 2018. Elevated Ictal Brain Network Ictogenicity Enables Prediction of Optimal Seizure Control. Frontiers in Neurology 9.

Martinet, L.-E., Fiddyment, G., Madsen, J. R., Eskandar, E. N., Truccolo, W., Eden, U. T., Cash, S. S., Kramer, M. A., Apr. 2017. Human seizures couple across spatial scales through travelling wave dynamics. Nature Communications 8 (1), 14896.

Medina Villalon, S., Paz, R., Roehri, N., Lagarde, S., Pizzo, F., Colombet, B., Bartolomei, F., Carron, R., Bénar, C.-G., Jun. 2018. EpiTools, A software suite for presurgical brain mapping in epilepsy: Intracerebral EEG. Journal of Neuroscience Methods 303, 7–15.

Milton, J. G., Chkhenkeli, S. A., Towle, V. L., 2007. Brain Connectivity and the Spread of Epileptic Seizures. In: Jirsa, V. K., McIntosh, A. (Eds.), Handbook of Brain Connectivity. Springer Berlin Heidelberg, Berlin, Heidelberg, pp. 477–503.

Murta, T., Leal, A., Garrido, M. I., Figueiredo, P., Sep. 2012. Dynamic Causal Modelling of epileptic seizure propagation pathways: A combined EEG–fMRI study. NeuroImage 62 (3), 1634–1642.

Neal, R., 2011. MCMC using Hamiltonian dynamics. Chapman and Hall/CRC.

Olmi, S., Petkoski, S., Guye, M., Bartolomei, F., Jirsa, V., Feb. 2019. Controlling seizure propagation in large-scale brain networks. PLOS Computational Biology 15 (2), e1006805.

Papadopoulou, M., Cooray, G., Rosch, R., Moran, R., Marinazzo, D., Friston, K., Feb. 2017. Dynamic causal modelling of seizure activity in a rat model. NeuroImage 146, 518–532.

Papadopoulou, M., Leite, M., van Mierlo, P., Vonck, K., Lemieux, L., Friston, K., Marinazzo, D., Feb. 2015. Tracking slow modulations in synaptic gain using dynamic causal modelling: Validation in epilepsy. NeuroImage 107, 117–126.

Parker, C. S., Clayden, J. D., Cardoso, M. J., Rodionov, R., Duncan, J. S., Scott, C., Diehl, B., Ourselin, S., 2018. Structural and effective connectivity in focal epilepsy. NeuroImage: Clinical 17, 943–952.

Parvizi, J., Kastner, S., Mar. 2018. Promises and limitations of human intracranial electroencephalography. Nature Neuroscience 21 (4), 474–483.

Perucca, P., Dubeau, F., Gotman, J., Oct. 2014. Intracranial electroencephalographic seizure-onset patterns: Effect of underlying pathology. Brain 137 (1), 183–196.

Proix, T., Bartolomei, F., Guye, M., Jirsa, V. K., Feb. 2017. Individual brain structure and modelling predict seizure propagation. Brain 140 (3), 641–654.

Proix, T., Spiegler, A., Schirner, M., Rothmeier, S., Ritter, P., Jirsa, V. K., Nov. 2016. How do parcellation size and short-range connectivity affect dynamics in large-scale brain network models? NeuroImage 142, 135–149.

Rosch, R. E., Hunter, P. R., Baldeweg, T., Friston, K. J., Meyer, M. P., Aug. 2018. Calcium imaging and dynamic causal modelling reveal brain-wide changes in effective connectivity and synaptic dynamics during epileptic seizures. PLOS Computational Biology 14 (8), e1006375.

Rossi, L. F., Wykes, R. C., Kullmann, D. M., Carandini, M., Aug. 2017. Focal cortical seizures start as standing waves and propagate respecting homotopic connectivity. Nature Communications 8 (1).

Scheffer, I. E., Berkovic, S., Capovilla, G., Connolly, M. B., French, J., Guilhoto, L., Hirsch, E., Jain, S., Mathern, G. W., Moshé, S. L., Nordli, D. R., Perucca, E., Tomson, T., Wiebe, S., Zhang, Y.-H., Zuberi, S. M., Apr. 2017. ILAE classification of the epilepsies: Position paper of the ILAE Commission for Classification and Terminology. Epilepsia 58 (4), 512–521.

Schevon, C. A., Weiss, S. A., McKhann, G., Goodman, R. R., Yuste, R., Emerson, R. G., Trevelyan, A. J., Jan. 2012. Evidence of an inhibitory restraint of seizure activity in humans. Nature Communications 3 (1).

Schroeder, G. M., Diehl, B., Chowdhury, F. A., Duncan, J. S., de Tisi, J., Trevelyan, A. J., Forsyth, R., Jackson, A., Taylor, P. N., Wang, Y., Jun. 2019. Seizure pathways change on circadian and slower timescales in individual patients with focal epilepsy. Preprint, Neuroscience.

Sevy, A., Gavaret, M., Trebuchon, A., Vaugier, L., Wendling, F., Carron, R., Regis, J., Chauvel, P., Gonigal, A. M., Bartolomei, F., May 2014. Beyond the lesion: The epileptogenic networks around cavernous angiomas. Epilepsy Research 108 (4), 701–708.

Sinha, N., Dauwels, J., Kaiser, M., Cash, S. S., Brandon Westover, M., Wang, Y., Taylor, P. N., Feb. 2017. Predicting neurosurgical outcomes in focal epilepsy patients using computational modelling. Brain 140 (2), 319–332.

Smith, E. H., Liou, J.-y., Davis, T. S., Merricks, E. M., Kellis, S. S., Weiss, S. A., Greger, B., House, P. A., Ii, G. M. M., Goodman, R. R., Emerson, R. G., Bateman, L. M., Trevelyan, A. J., Schevon, C. A., Mar. 2016. The ictal wavefront is the spatiotemporal source of discharges during spontaneous human seizures. Nature Communications 7, 11098.

Taylor, P. N., Kaiser, M., Dauwels, J., Oct. 2014. Structural connectivity based whole brain modelling in epilepsy. Journal of Neuroscience Methods 236, 51–57.

Terry, J. R., Benjamin, O., Richardson, M. P., 2012. Seizure generation: The role of nodes and networks. Epilepsia 53 (9), e166–e169.

Tournier, J.-D., Calamante, F., Connelly, A., May 2007. Robust determination of the fibre orientation distribution in diffusion MRI: Non-negativity constrained super-resolved spherical deconvolution. NeuroImage 35 (4), 1459–1472.

Tournier, J.-D., Calamante, F., Connelly, A., 2010. Improved probabilistic streamlines tractography by 2nd order integration over fibre orientation distributions. In: Proceedings of the International Society for Magnetic Resonance in Medicine. Vol. 18. p. 1670.

Tournier, J.-D., Calamante, F., Connelly, A., Aug. 2013. Determination of the appropriate b-value and number of gradient directions for high-angular-resolution diffusion-weighted imaging. NMR in Biomedicine 26 (12), 1775–1786.

Wagner, F. B., Eskandar, E. N., Cosgrove, G. R., Madsen, J. R., Blum, A. S., Potter, N. S., Hochberg, L. R., Cash, S. S., Truccolo, W., Nov. 2015. Microscale spatiotemporal dynamics during neocortical propagation of human focal seizures. NeuroImage 122, 114–130.

Wang, Y., Trevelyan, A. J., Valentin, A., Alarcon, G., Taylor, P. N., Kaiser, M., May 2017. Mechanisms underlying different onset patterns of focal seizures. PLOS Computational Biology 13 (5), e1005475.

Wendling, F., Benquet, P., Bartolomei, F., Jirsa, V., Feb. 2016. Computational models of epileptiform activity. Journal of Neuroscience Methods 260, 233–251.

Wirsich, J., Perry, A., Ridley, B., Proix, T., Golos, M., Bénar, C., Ranjeva, J.-P., Bartolomei, F., Breakspear, M., Jirsa, V., Guye, M., Jan. 2016. Whole-brain analytic measures of network communication reveal increased structure-function correlation in right temporal lobe epilepsy. NeuroImage: Clinical 11, 707–718.

## References

Betancourt, M., Mar. 2018. Calibrating Model-Based Inferences and Decisions.

